# Pediatric outpatient visits and antibiotic use attributable to higher valency pneumococcal conjugate vaccine serotypes

**DOI:** 10.1101/2023.08.24.23294570

**Authors:** Laura M King, Kristin L Andrejko, Sarah Kabbani, Sara Y Tartof, Lauri A Hicks, Adam L Cohen, Miwako Kobayashi, Joseph A Lewnard

**Author notes:** **Corresponding author:** Laura M King, MPH, University of California, School of Public Health, 2121 Berkeley Way, Berkeley, CA 94704. Co-senior authors (Miwako Kobayashi, Joseph A Lewnard).

## Abstract

**Importance:** *Streptococcus pneumoniae* is a known etiology of acute respiratory infections (ARIs), which account for large proportions of outpatient visits and antibiotic use in children. In 2023, 15- and 20-valent pneumococcal conjugate vaccines (PCV15, PCV20) were recommended for routine use in infants. However, the burden of outpatient healthcare utilization among U.S. children attributable to the additional, non-PCV13 serotypes in PCV15/20 is unknown.

**Objective:** To estimate the incidence of outpatient visits and antibiotic prescriptions in U.S. children for acute otitis media, pneumonia, and sinusitis associated with PCV15- and PCV20-additional serotypes (non-PCV13 serotypes) to quantify potential impacts of PCV15/20 on outpatient visits and antibiotic prescriptions for these conditions.

**Design:** Multi-component study including descriptive analyses of cross-sectional and cohort data on outpatient visits and antibiotic prescriptions from 2016–2019 and meta-analyses of pneumococcal serotype distribution in non-invasive respiratory infections.

**Setting:** Outpatient visits and antibiotic prescriptions among U.S. children.

**Participants:** Pediatric visits and antibiotic prescriptions among children captured in the National Ambulatory Medical Care Survey (NAMCS), the National Hospital Ambulatory Medicare Care Survey (NHAMCS), and Merative MarketScan, collectively representing healthcare delivery across all outpatient settings. Incidence denominators estimated using census (NAMCS/NHAMCS) and enrollment (MarketScan) data.

**Main outcome(s) and measure(s):** Pediatric outpatient visit and antibiotic prescription incidence for acute otitis media, pneumonia, and sinusitis associated with PCV15/20-additional serotypes.

**Results:** We estimated that per 1000 children annually, PCV15-additional serotypes accounted for 2.7 (95% confidence interval 1.8–3.9) visits and 2.4 (1.6–3.4) antibiotic prescriptions. PCV20-additional serotypes resulted in 15.0 (11.2–20.4) visits and 13.2 (9.9–18.0) antibiotic prescriptions annually per 1,000 children. Projected to national counts, PCV15/20-additional serotypes account for 173,000 (118,000–252,000) and 968,000 (722,000–1,318,000) antibiotic prescriptions among U.S. children each year, translating to 0.4% (0.2–0.6%) and 2.1% (1.5–3.0%) of all outpatient antibiotic use among children.

**Conclusions and relevance:** PCV15/20-additional serotypes account for a large burden of pediatric outpatient healthcare utilization. Compared with PCV15-additional serotypes, PCV20-additional serotypes account for >5 times the burden of visits and antibiotic prescriptions. These higher-valency PCVs, especially PCV20, may contribute to preventing ARIs and antibiotic use in children.

**Key points:** **Question:** What is the incidence of pediatric outpatient visits and antibiotic prescriptions for acute respiratory tract infections associated with the additional (non-13-valent pneumococcal conjugate vaccine) serotypes in 15- and 20-valent pneumococcal conjugate vaccines (PCV15, PCV20)?

**Findings:** PCV15- and PCV20-additional serotypes account for an estimated 197,000 and 1,098,000 visits and 173,000 and 968,000 antibiotic prescriptions, respectively, for acute respiratory infections among U.S. children annually. Visit and antibiotic prescription burdens were concentrated among children aged <5 years.

**Meaning:** Respiratory infections caused by serotypes in PCV15/20, especially PCV20, may be important contributors to outpatient visits and antibiotic use in children.

## Introduction

Acute respiratory infections (ARIs) are a leading cause of outpatient healthcare utilization,^1^ school absenteeism,^2^ and antibiotic use^3,4^ among children, resulting in considerable health and economic burdens. In 2014–15, ARIs accounted for 544 outpatient visits and 252 antibiotic prescriptions per 1000 children annually, with acute otitis media (AOM) accounting for over a third of ARI-associated antibiotic prescriptions.^5^ *Streptococcus pneumoniae* (pneumococcus) causes bacterial ARIs among children, especially AOM and thus contributes to antibiotic use in children. Pneumococcal conjugate vaccines (PCVs) provide protection against carriage of, and disease from, vaccine-targeted serotypes. Implementation of PCV7, followed by PCV13 in 2011,^6^ contributed to substantial reductions in AOM and pneumonia incidence among children in the United States.^7–16^

In 2023, the Advisory Committee on Immunization Practices (ACIP) amended pediatric pneumococcal vaccine recommendations to include PCV15 or PCV20 in a series of 3 primary doses (ages 2, 4, and 6 months) and one booster dose (12–15 months).^17,18^ These vaccines extend PCV13 serotype composition (serotypes 1, 3, 4, 5, 6A, 6B, 7A, 9V, 14, 18C, 19A, 19F, 23F) to include 22F and 33F (PCV15/20) and 8, 10A, 11A, 12F, and 15B (PCV20). Twenty-three-valent pneumococcal polysaccharide vaccine (PPSV23) is recommended only for children aged >2 years with certain comorbidities and previous PCV13/15 immunization.^18^

Serotypes included in PCV15 and PCV20 accounted for 31% and 44%, respectively, of invasive pneumococcal disease (IPD) cases among U.S. children <5 years in 2020–21^19^ and PCVs have been formulated to include serotypes causing IPD.^20^ Pediatric IPD incidence has reached historic lows, with 7.2 cases per 100,000 children aged <5 years in 2019.^21^ Less is known about the burden of outpatient ARIs attributable to PCV15/20 serotypes. However, consideration of the burden of outpatient outcomes in the development of vaccine recommendations and evaluation of vaccine implementation is warranted due to the high burden of outpatient outcomes,^4,5,9,16,22^, their role in vaccine cost-effectiveness,^23,24^ and needed reductions in antibiotic use.^25^ We therefore aimed to estimate the incidence of outpatient visits and antibiotic prescriptions in children in the United States for AOM, pneumonia, and sinusitis associated with the additional (non-PCV13) serotypes in PCV15 and PCV20.

## Methods

### Estimation framework

Our approach to estimate the incidence of pediatric outpatient visits and antibiotic prescriptions attributable to additional serotypes in PCV15 and PCV20 (hereafter, PCV15/20-additional serotypes) included three main components (**Figure 1**). First, we estimated all-cause incidence rates of outpatient visits and antibiotic prescriptions for AOM, pneumonia, and sinusitis. Second, we estimated the proportion of cases of these conditions attributable to PCV15/20-additional serotypes. Finally, we obtained incidence rates of visits and antibiotic prescriptions attributable to PCV15/20-additional serotypes by multiplying the all-cause incidence rates and corresponding attributable proportion estimates obtained in the previous steps.

**Figure 1.**
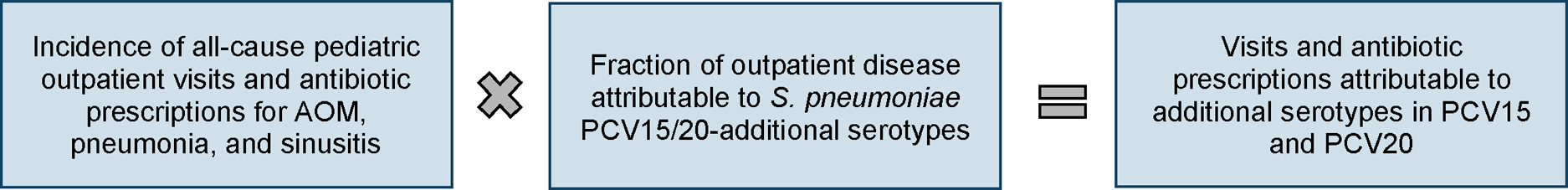
Estimation framework. Framework to derive estimates of visits and antibiotic prescriptions attributable to pneumococcal serotypes included in 15- and 20-valent pneumococcal conjugate vaccines (PCV15/20) not included in PCV13. All-cause incidence estimation methods are detailed in **eMethods 1**. Primary attributable fraction analysis methods are detailed in **eMethods 2** (AOM) and **eMethods 3** (pneumonia, sinusitis) and supplemental analysis methods are detailed in **eMethods 5.**

We limited our study to children because children are targeted for universal PCV immunization and account for high proportions of outpatient visits and antibiotic prescriptions for ARIs.^4,5^ We generated estimates for all children and age-specific strata (<2 years, 2–4 years, 5–17 years). For age stratum estimates, we only considered sinusitis in children aged 5–17 years as full sinus development typically occurs >5 years of age^26,27^ and sinusitis is infrequently diagnosed in younger children. Estimates for all children (0–17 years) include all three conditions.

To contextualize estimated burdens, we projected incidence per 1000 person-years to national numbers of visits and antibiotic prescriptions by multiplying estimated incidence rates by corresponding 2019 bridged-race postcensal population estimates (Vintage 2020).^28^ Additionally, we estimated the proportion of all pediatric antibiotic prescribing attributable to PCV15/20-additional serotypes by dividing estimated PCV15/20-additional serotype-attributable incidence rates by the incidence of all pediatric outpatient antibiotic prescriptions.

### All-cause visit and antibiotic prescription incidence rates

We estimated incidence rates of visits and antibiotic prescriptions for AOM, pneumonia, and sinusitis among children aged <18 years across U.S. outpatient settings using the 2016 and 2019 National Ambulatory Medical Care Survey (NAMCS), 2016 and 2019 National Hospital Ambulatory Medicare Care Survey (NHAMCS), and 2016–2019 Merative MarketScan Commercial and Medicaid Databases (MarketScan), as detailed in **eMethods 1**. Collectively, these datasets represent healthcare delivery across all outpatient settings (**eTable 1**). We used NAMCS and NHAMCS to obtain nationally representative incidence estimates from physician offices and emergency departments. We generated incidence estimates from other outpatient settings (e.g., urgent care, retail health facilities) using data from MarketScan, which contains reconciled claims from all outpatient settings among a large convenience sample of individuals with commercial insurance and Medicaid. Total MarketScan rates were estimated as weighted averages from the Commercial and Medicaid databases with weights accounting for distributions of commercial and public insurance (**eMethods 1, eTable 2**). We identified visits and prescriptions for AOM, pneumonia, and sinusitis from *International Classification of Diseases, Tenth Revision, Clinical Modification* (ICD-10-CM) diagnosis codes using a previously validated algorithm that assigns a single diagnosis to each visit based on the diagnosis code most likely to result in an antibiotic prescription (**eMethods 1**, **eTable 3**).^3–5^ To estimate incidence rates across all outpatient settings, we standardized estimates from each data source per 1000 population and summed the rates from office and emergency departments (NAMCS/NHAMCS) and all other outpatient settings (MarketScan). This activity was reviewed by CDC and was conducted consistent with applicable federal law and CDC policy.§ (§See e.g., 45 C.F.R. part 46, 21 C.F.R. part 56; 42 U.S.C. §241(d); 5 U.S.C. §552a; 44 U.S.C. §3501 et seq.).

### Serotype-specific attributable fractions

To estimate pneumococcal serotype-specific attributable fractions, we used a modified vaccine-probe approach comparing observed changes in incidence of each condition following PCV13 implementation (**eMethods 2–3)**. In vaccine-probe studies, the proportion of disease attributable to a vaccine-targeted pathogen is estimated by dividing vaccine effectiveness (VE) against all-cause disease by VE against the vaccine-targeted pathogen.^29^ Analyses accounted for secular trends in incidence using “negative-control” conditions not likely to have been impacted by PCVs (**eTable 4**). We projected PCV13 estimates from the vaccine probe analysis to PCV15/20 serotypes using the ratio of PCV13 versus PCV15/20 serotype frequencies in the studied ARI conditions, estimated through meta-analyses of published estimates of serotype distributions (**eMethods 4, eTable 5**). We categorized serotypes as PCV13, PCV15-additional, and PCV20-additional as described in **eTable 6**. As PPSV23 is only recommended in select children, we did not consider PPSV23 serotypes in this analysis. Studies in older children were limited; we assumed serotype distribution was consistent across age groups.

Etiology data sources for this analysis framework differed across the three ARI conditions. For AOM, microbiological surveillance data were available from Israeli children aged <3 years from 2009–11 and 2013– 15,^30^ allowing us to directly compare changes in all-cause and PCV13-serotype AOM. Similar to the United States, PCV13 replaced PCV7 in Israel in 2010.^31,32^ We used these data to estimate vaccine effectiveness (VE) against all-cause and PCV13 serotype AOM, with non-pneumococcal AOM as a negative-control (**eMethods 2, eTable 4**). As similar, microbiologically-detailed studies of pneumonia and sinusitis etiology were not available, we estimated VE against all-cause pneumonia and sinusitis in children <3 years from 2009–10 and 2013–15 using NAMCS/NHAMCS, with skin and soft tissue infections as a negative-control (**eMethods 3, eTable 4**). We assumed VE against PCV13 serotype disease was equivalent for these conditions as for AOM.

### Attributable fraction sensitivity analyses

We supplemented this analysis using two additional attributable fraction estimation methods (**eMethods 5**). First, we estimated the difference in pneumococcal nasopharyngeal carriage prevalence among children with ARI/AOM and healthy children and defined this prevalence difference as the proportion of cases attributable to pneumococci (differential carriage approach). This approach was grounded in findings from previous studies demonstrating increased pneumococcal carriage prevalence during ARI episodes.^33,34^ We undertook a systematic literature review and meta-analysis to determine pneumococcal carriage prevalence among healthy children and those with AOM/ARIs (**eMethods 5, eTable 7**) and used our previously-estimated nasopharyngeal serotype distributions (**eMethods 4, eTable 5**). We conducted this analysis for AOM only and all ARIs. Second, for AOM, we also undertook a meta-analysis estimating attributable fractions using published estimates of pneumococcal prevalence and serotype distribution in middle ear fluid (MEF) samples from children with AOM (**eMethods 5; eTable 8**).

We undertook additional sensitivity analyses to evaluate the impact of study selection on our findings. First, we restricted included studies of serotype distributions to those undertaken ≥4 years post-PCV13 implementation (**eMethods 6**). In the second, we excluded published isolate counts that were aggregated across multiple serotypes except 15B/C (**eMethods 4**).

### Total burden due to PCV15/20 serotypes

Finally, to quantify the total serotype coverage of these vaccines, we estimated incidence attributable to all serotypes in PCV15 and PCV20 including disease from PCV15/20-additional serotypes, residual PCV13 disease not prevented by PCV13, and disease prevented by PCV13 during the study period (**eMethods 2–3**). This analysis was based on a counterfactual of no PCV use (distinct from the continued PCV13 use counterfactual in primary analyses), as the total benefit of these vaccines includes continuation of PCV13 serotype protection.^35^

## Results

### All-cause AOM, pneumonia, and sinusitis incidence

For all conditions, there were 234 outpatient visits (95% confidence interval: 193–281) and 206 (173–243) outpatient antibiotic prescriptions per 1000 person-years among U.S. children during 2016–2019 (**Table 1**; **eTable 2**), projecting to 17.1 (14.1–20.5) million outpatient visits and 15.0 (12.7–17.7) million antibiotic prescriptions annually. The highest incidence rates of visits and prescriptions were observed among children aged <5 years. By condition, AOM accounted for the highest incidence rates, with 166 (130–209) visits and 144 (118–174) antibiotic prescriptions per 1000 person-years among all children. In children aged <5 years, AOM accounted for 451 (353–567) visits and 365 (288–455) antibiotic prescriptions per 1000 person-years. There were 17 (12–23) visits and 17 (13–21) antibiotic prescriptions per 1000 person-years for pneumonia and 49 (33–72) visits and 44 (27–67) antibiotic prescriptions for sinusitis among children 0–17 years.

**Table 1.**
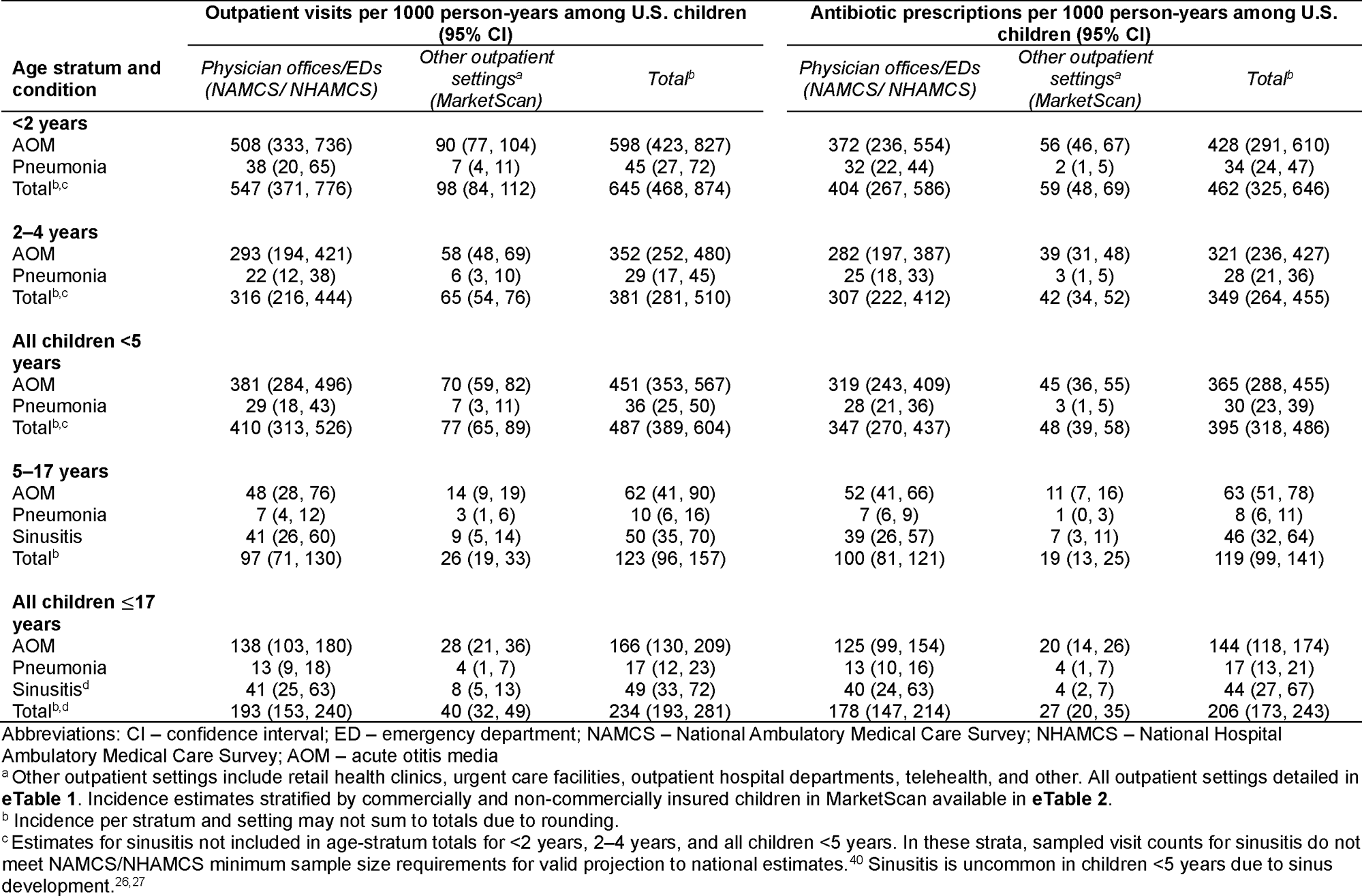
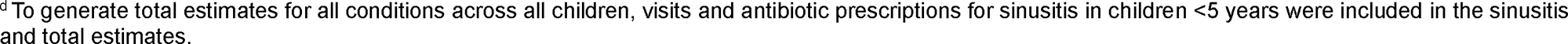
Estimated incidence of all-cause pediatric visits and antibiotic prescriptions by outpatient setting and data source, 2016–2019.

| Age stratum and condition | Outpatient visits per 1000 person-years among U.S. children (95% CI) |  |  | Antibiotic prescriptions per 1000 person-years among U.S. children (95% CI) |  |  |
| --- | --- | --- | --- | --- | --- | --- |
|  | Physician offices/EDs (NAMCS/ NHAMCS) | Other outpatient settings<sup>a</sup> (MarketScan) | Total<sup>b</sup> | Physician offices/EDs (NAMCS/ NHAMCS) | Other outpatient settings<sup>a</sup> (MarketScan) | Total<sup>b</sup> |
| **<2 years** |  |  |  |  |  |  |
| AOM | 508 (333, 736) | 90 (77, 104) | 598 (423, 827) | 372 (236, 554) | 56 (46, 67) | 428 (291, 610) |
| Pneumonia | 38 (20, 65) | 7 (4, 11) | 45 (27, 72) | 32 (22, 44) | 2 (1, 5) | 34 (24, 47) |
| Total<sup>b,c</sup> | 547 (371, 776) | 98 (84, 112) | 645 (468, 874) | 404 (267, 586) | 59 (48, 69) | 462 (325, 646) |
| **2–4 years** |  |  |  |  |  |  |
| AOM | 293 (194, 421) | 58 (48, 69) | 352 (252, 480) | 282 (197, 387) | 39 (31, 48) | 321 (236, 427) |
| Pneumonia | 22 (12, 38) | 6 (3, 10) | 29 (17, 45) | 25 (18, 33) | 3 (1, 5) | 28 (21, 36) |
| Total<sup>b,c</sup> | 316 (216, 444) | 65 (54, 76) | 381 (281, 510) | 307 (222, 412) | 42 (34, 52) | 349 (264, 455) |
| **All children <5 years** |  |  |  |  |  |  |
| AOM | 381 (284, 496) | 70 (59, 82) | 451 (353, 567) | 319 (243, 409) | 45 (36, 55) | 365 (288, 455) |
| Pneumonia | 29 (18, 43) | 7 (3, 11) | 36 (25, 50) | 28 (21, 36) | 3 (1, 5) | 30 (23, 39) |
| Total<sup>b,c</sup> | 410 (313, 526) | 77 (65, 89) | 487 (389, 604) | 347 (270, 437) | 48 (39, 58) | 395 (318, 486) |
| **5–17 years** |  |  |  |  |  |  |
| AOM | 48 (28, 76) | 14 (9, 19) | 62 (41, 90) | 52 (41, 66) | 11 (7, 16) | 63 (51, 78) |
| Pneumonia | 7 (4, 12) | 3 (1, 6) | 10 (6, 16) | 7 (6, 9) | 1 (0, 3) | 8 (6, 11) |
| Sinusitis | 41 (26, 60) | 9 (5, 14) | 50 (35, 70) | 39 (26, 57) | 7 (3, 11) | 46 (32, 64) |
| Total<sup>b</sup> | 97 (71, 130) | 26 (19, 33) | 123 (96, 157) | 100 (81, 121) | 19 (13, 25) | 119 (99, 141) |
| **All children ≤17 years** |  |  |  |  |  |  |
| AOM | 138 (103, 180) | 28 (21, 36) | 166 (130, 209) | 125 (99, 154) | 20 (14, 26) | 144 (118, 174) |
| Pneumonia | 13 (9, 18) | 4 (1, 7) | 17 (12, 23) | 13 (10, 16) | 4 (1, 7) | 17 (13, 21) |
| Sinusitis<sup>d</sup> | 41 (25, 63) | 8 (5, 13) | 49 (33, 72) | 40 (24, 63) | 4 (2, 7) | 44 (27, 67) |
| Total<sup>b,d</sup> | 193 (153, 240) | 40 (32, 49) | 234 (193, 281) | 178 (147, 214) | 27 (20, 35) | 206 (173, 243) |

Abbreviations: CI – confidence interval; ED – emergency department; NAMCS – National Ambulatory Medical Care Survey; NHAMCS – National Hospital Ambulatory Medical Care Survey; AOM – acute otitis media
<sup>a</sup> Other outpatient settings include retail health clinics, urgent care facilities, outpatient hospital departments, telehealth, and other. All outpatient settings detailed in **eTable 1**. Incidence estimates stratified by commercially and non-commercially insured children in MarketScan available in **eTable 2**.
<sup>b</sup> Incidence per stratum and setting may not sum to totals due to rounding.
<sup>c</sup> Estimates for sinusitis not included in age-stratum totals for <2 years, 2–4 years, and all children <5 years. In these strata, sampled visit counts for sinusitis do not meet NAMCS/NHAMCS minimum sample size requirements for valid projection to national estimates.<sup>40</sup> Sinusitis is uncommon in children <5 years due to sinus development.<sup>26,27</sup>
<sup>d</sup>To generate total estimates for all conditions across all children, visits and antibiotic prescriptions for sinusitis in children <5 years were included in the sinusitis and total estimates.

### Pneumococcal serotype distribution in ARIs with pneumococcal detection

Data on pneumococcal serotype distributions in nasopharyngeal carriage among children experiencing ARI were available from five studies; four enrolled children with AOM, and one enrolled children with lower respiratory tract infection (**eTable 5**). Across these studies, the most common serotypes were 19A, 3, 15B/C, 35B, and 11A (**Figure 2**). Additional serotypes in PCV15 and PCV20 accounted for 3.6% (2.8–4.4%) and 22.7% (21.0–24.5%), respectively, of pneumococcal isolates from nasopharyngeal samples in ARIs (**eTable 9**). In AOM, PCV15-additional and PCV20-additional serotypes accounted for 5.2% (3.7–7.1%) and 26.9% (23.5– 30.4%) of pneumococcal isolates. Non-vaccine serotypes accounted for 48.6% (44.6–52.5%) of isolates in children with AOM and 54.0% (51.9–56.1%) of isolates in children with ARI.

**Figure 2.**
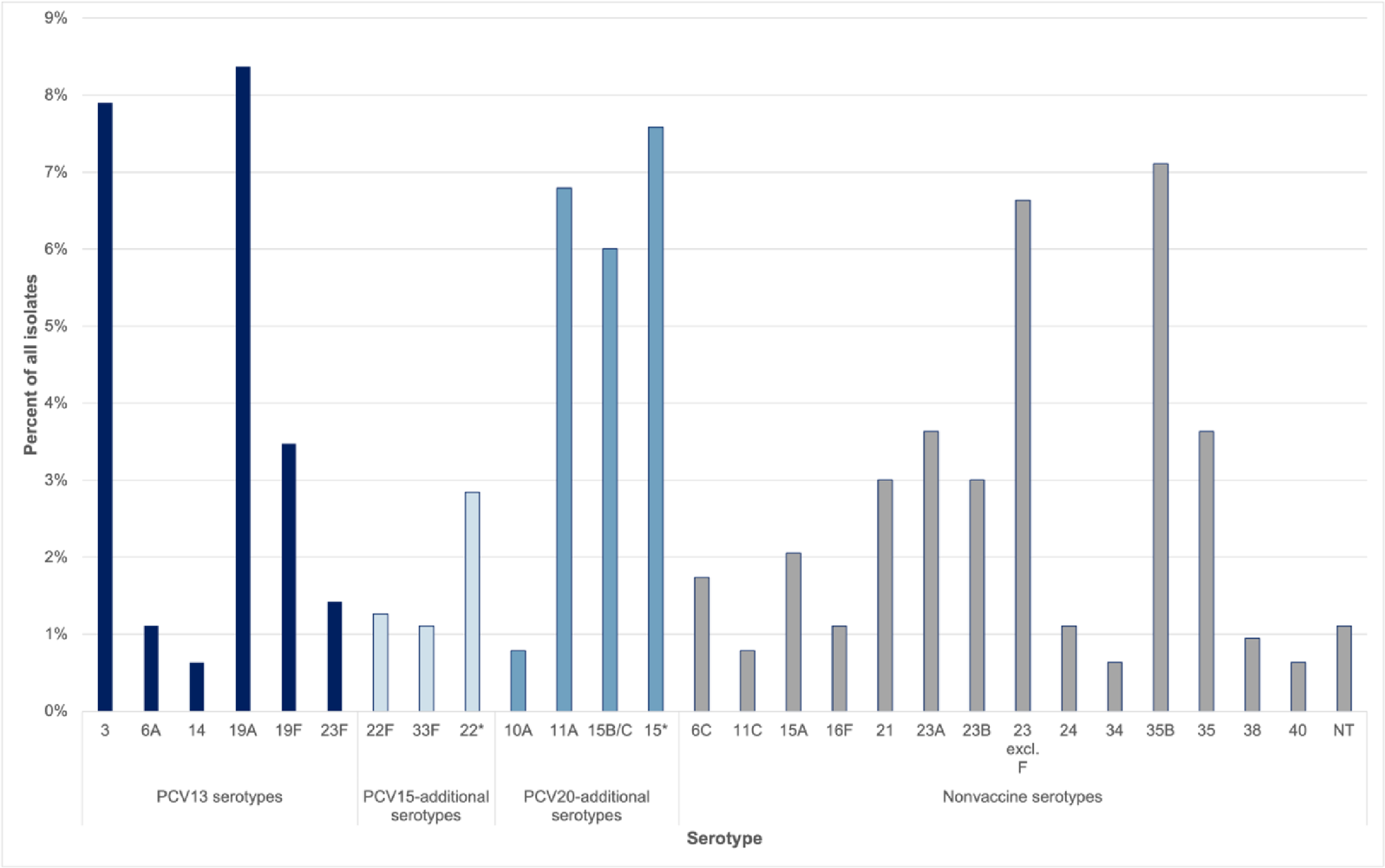
Distribution of PCV13, PCV15-additional, and PCV20-additional serotypes among children with outpatient ARIs. We illustrate the proportions of isolates belonging to serotypes across studies included in the meta-analysis of pneumococcal serotype distributions in ARIs (**eMethods 4**; **eTable 5**) by serotype and PCV inclusion category (PCV13: serotypes 1, 3, 4, 5, 6A, 6B, 7A, 9V, 14, 18C, 19A, 19F, 23F; PCV15-additional: 22F, 33F; PCV20-additional: 22F, 33F, 8, 10A, 11A, 12F, 15B). Only serotypes representing ≥1% of isolates presented. Related aggregated serotypes denoted by (*); see **eTable 6** for detailed serotype categorization. Data presented here are pooled directly from studies in **eTable 5;** Markov chain Monte Carlo estimates by PCV inclusion category are presented in **eTable 9**.

### Fraction of ARIs attributable to PCV15/20-additional serotypes

Using the vaccine probe method, we estimated that PCV15-additional serotypes accounted for 1.0% (0.7– 1.5%), 1.0% (0.6–1.5%), and 1.6% (1.1–2.6%) of current all-cause AOM, pneumonia, and sinusitis, respectively (**eTable 10**). PCV20-additional serotypes accounted for 5.2% (4.2–6.5%), 6.2% (4.2–9.3%), and 10.4% (7.1–15.5%) of all-cause AOM, pneumonia, and sinusitis. Estimates of PCV15/20 serotype coverage in AOM from alternative approaches closely resembled those from our primary vaccine-probe analyses. However, the differential carriage approach yielded lower estimates of the fraction of pneumonia and sinusitis cases attributable to pneumococci. Under this framework, PCV15-additional serotypes accounted for 0.4% (0.0– 0.8%) of all-cause disease, while PCV20-additional serotypes accounted for 2.7% (0.2–5.2%).

### Burden due to PCV15/20-additional serotypes

Annually per 1,000 children, PCV15-additional serotypes accounted for 2.7 (1.8–3.9) ARI visits and 2.4 (1.6– 3.4) antibiotic prescriptions (**Tables 2–3**). Projecting these rates to all U.S. children yielded 197,000 (133,000– 287,000) visits and 173,000 (118,000–252,000) antibiotic prescriptions attributable to PCV15-additional serotypes annually (**eTables 11–12**). In contrast, PCV20-additional serotypes resulted in 15.0 (11.2–20.4) ARI visits and 13.2 (9.9–18.0) antibiotic prescriptions annually per 1000 children, corresponding to 1,098,000 (816,000–1,489,000) visits and 968,000 (722,000–1,318,000) antibiotic prescriptions. In total, PCV15- and PCV20-additional serotypes accounted for 0.4% (0.2–0.6%) and 2.1% (1.5–3.0%), respectively, of antibiotic use among U.S. children (**eTable 13**).

**Table 2.** Outpatient visits attributable to any pneumococcal serotype, PCV15-additional serotypes, and PCV20-additional serotypes, 2016–2019.

| Age stratum and condition | Estimated incidence per 1000 person-years among U.S. children (95% CI) |  |  |
| --- | --- | --- | --- |
|  | *All pneumococci* | *PCV15-additional serotypes<sup>a</sup>* | *PCV20-additional serotypes<sup>a</sup>* |
| **<2 years** |  |  |  |
| AOM | 116.5 (80.3, 165.9) | 6.1 (3.6, 10.0) | 31.2 (20.8, 46.3) |
| Pneumonia | 12.3 (6.5, 22.7) | 0.4 (0.2, 0.8) | 2.8 (1.5, 5.2) |
| Total<sup>b,c</sup> | 129.3 (92.2, 179.8) | 6.5 (4.0, 10.5) | 34.2 (23.4, 49.5) |
| **2–4 years** |  |  |  |
| AOM | 68.4 (47.7, 96.2) | 3.6 (2.1, 5.8) | 18.3 (12.3, 26.9) |
| Pneumonia | 7.9 (4.2, 14.3) | 0.3 (0.1, 0.5) | 1.8 (0.9, 3.3) |
| Total<sup>b,c</sup> | 76.6 (55.3, 105.2) | 3.9 (2.4, 6.1) | 20.2 (14.0, 29.0) |
| **All children <5 years** |  |  |  |
| AOM | 87.6 (66.4, 114.8) | 4.6 (2.9, 7.1) | 23.5 (17.0, 32.4) |
| Pneumonia | 9.7 (5.8, 16.3) | 0.3 (0.2, 0.6) | 2.2 (1.3, 3.7) |
| Total<sup>b,c</sup> | 97.7 (75.7, 125.8) | 4.9 (3.2, 7.5) | 25.8 (19.0, 34.8) |
| **5–17 years** |  |  |  |
| AOM | 12.1 (7.9, 18.0) | 0.6 (0.4, 1.1) | 3.2 (2.1, 5.0) |
| Pneumonia | 2.7 (1.4, 5.0) | 0.1 (0.0, 0.2) | 0.6 (0.3, 1.2) |
| Sinusitis | 22.9 (13.7, 38.3) | 0.8 (0.5, 1.4) | 5.2 (3.1, 8.8) |
| Total<sup>b</sup> | 38.2 (26.7, 55.5) | 1.6 (1.0, 2.4) | 9.2 (6.4, 13.4) |
| **All children ≤17 years** |  |  |  |
| AOM | 32.3 (24.5, 42.3) | 1.7 (1.1, 2.6) | 8.7 (6.3, 11.9) |
| Pneumonia | 4.6 (2.8, 7.6) | 0.2 (0.1, 0.3) | 1.1 (0.6, 1.7) |
| Sinusitis<sup>d</sup> | 22.6 (13.1, 38.9) | 0.8 (0.4, 1.4) | 5.1 (2.9, 8.9) |
| Total<sup>b,d</sup> | 60.1 (45.8, 80.5) | 2.7 (1.8, 3.9) | 15.0 (11.2, 20.4) |

Abbreviations: PCV – pneumococcal conjugate vaccine; AOM – acute otitis media
<sup>a</sup> Includes vaccine serotypes and corresponding aggregated serotypes/serogroups as detailed in **eTable 6**. PCV-15 additional serotypes (not included in PCV13): 22F, 33F. PCV20-additional serotypes: 8, 10A, 11A, 12F, 15B, 22F, 33F.
<sup>b</sup> Incidence by condition may not sum to total due to rounding.
<sup>c</sup> Estimates for sinusitis not included in age-stratum totals for <2 years, 2–4 years, and all children <5 years. In these strata, sampled visit counts for sinusitis do not meet NAMCS/NHAMCS minimum sample size requirements for valid projection to national estimates.<sup>40</sup> Sinusitis is uncommon in children <5 years due to sinus development.<sup>26,27</sup>
<sup>d</sup> To generate total estimates for all conditions across all children, visits and antibiotic prescriptions for sinusitis in children <5 years were included in the sinusitis and total estimates.

**Table 3.** Antibiotic prescriptions attributable to any pneumococcal serotype, PCV15-additional serotypes, and PCV20-additional serotypes, 2016–2019.

| Age stratum and condition | Estimated incidence per 1000 person-years among U.S. children (95% CI) |  |  |
| --- | --- | --- | --- |
|  | *All pneumococci* | *PCV15-additional serotypes<sup>a</sup>* | *PCV20-additional serotypes<sup>a</sup>* |
| **<2 years** |  |  |  |
| AOM | 83.2 (55.3, 122.0) | 4.3 (2.5, 7.3) | 22.3 (14.3, 34.0) |
| Pneumonia | 9.4 (5.7, 15.5) | 0.3 (0.2, 0.6) | 2.1 (1.3, 3.6) |
| Total<sup>b,c</sup> | 92.9 (64.4, 132.2) | 4.7 (2.8, 7.7) | 24.5 (16.4, 36.4) |
| **2–4 years** |  |  |  |
| AOM | 62.4 (44.7, 85.8) | 3.3 (2.0, 5.2) | 16.7 (11.5, 24.1) |
| Pneumonia | 7.6 (4.7, 12.1) | 0.3 (0.2, 0.5) | 1.7 (1.1, 2.8) |
| Total<sup>b,c</sup> | 70.2 (51.9, 94.0) | 3.5 (2.2, 5.5) | 18.5 (13.1, 26.0) |
| **All children <5 years** |  |  |  |
| AOM | 70.9 (54.0, 92.4) | 3.7 (2.4, 5.7) | 19.0 (13.8, 26.0) |
| Pneumonia | 8.3 (5.2, 13.2) | 0.3 (0.2, 0.5) | 1.9 (1.2, 3.0) |
| Total<sup>b,c</sup> | 79.5 (61.9, 101.7) | 4.0 (2.6, 6.0) | 21.0 (15.5, 28.1) |
| **5–17 years** |  |  |  |
| AOM | 12.3 (9.6, 15.8) | 0.6 (0.4, 1.0) | 3.3 (2.4, 4.5) |
| Pneumonia | 2.3 (1.5, 3.6) | 0.1 (0.0, 0.1) | 0.5 (0.3, 0.8) |
| Sinusitis | 21.2 (12.7, 35.2) | 0.8 (0.4, 1.3) | 4.8 (2.9, 8.1) |
| Total<sup>b</sup> | 36.0 (26.2, 51.3) | 1.5 (1.0, 2.2) | 8.7 (6.3, 12.4) |
| **All children ≤17 years** |  |  |  |
| AOM | 28.0 (22.0, 35.5) | 1.5 (0.9, 2.2) | 7.5 (5.6, 10.1) |
| Pneumonia | 4.6 (2.9, 7.2) | 0.2 (0.1, 0.3) | 1.0 (0.7, 1.6) |
| Sinusitis<sup>d</sup> | 20.2 (11.2, 35.7) | 0.7 (0.4, 1.3) | 4.6 (2.5, 8.2) |
| Total<sup>b,d</sup> | 53.1 (40.6, 71.8) | 2.4 (1.6, 3.4) | 13.2 (9.9, 18.0) |

Abbreviations: PCV – pneumococcal conjugate vaccine; AOM – acute otitis media
<sup>a</sup> Includes vaccine serotypes and corresponding aggregated serotypes/serogroups as detailed in **eTable 6**. PCV-15 additional serotypes (not included in PCV13): 22F, 33F. PCV20-additional serotypes: 8, 10A, 11A, 12F, 15B, 22F, 33F.
<sup>b</sup> Incidence by condition may not sum to total due to rounding.
<sup>c</sup> Estimates for sinusitis not included in age-stratum totals for <2 years, 2–4 years, and all children <5 years. In these strata, sampled visit counts for sinusitis do not meet NAMCS/NHAMCS minimum sample size requirements for valid projection to national estimates.<sup>40</sup> Sinusitis is uncommon in children <5 years due to sinus development.<sup>26,27</sup>
<sup>d</sup> To generate total estimates for all conditions across all children, visits and antibiotic prescriptions for sinusitis in children <5 years were included in the sinusitis and total estimates.

### Sensitivity analyses for attributable fractions

Compared with primary analyses, restricting analyses to include studies undertaken ≥4 years after PCV13 implementation yielded greater proportions of disease attributable to PCV15-additional and PCV20-additional serotypes (**eTables 14–15**), reflecting the enhanced contribution of these serotypes to all-cause ARIs from serotype replacement and declines in PCV13 serotype disease following PCV13 implementation. Compared with primary analyses, lower proportions of disease attributable to PCV20-additional serotypes were observed when analyses included only counts disaggregated by serogroup/serotype (**eTables 16–17**), likely due to the exclusion of samples categorized as serogroup 15.

### Total burden due to PCV15 and PCV20 serotypes

Considering disease associated with PCV15/20-additional serotypes and PCV13 serotypes (prevented by PCV13 and residual), we estimate all PCV15 serotypes would account for 50.9 (22.6–91.6) outpatient visits and 45.0 (20.2–79.6) antibiotic prescriptions per 1,000 person-years, while all PCV20 serotypes would account for 62.4 (32.0–106.8) outpatient visits and 55.1 (28.5–93.0) antibiotic prescriptions per 1,000 person-years (eTables 18–19).

## Discussion

We estimate that additional serotypes contained in PCV15 (22F, 33F) account for approximately 197,000 outpatient visits and 173,00 outpatient antibiotic prescriptions for AOM, pneumonia, and sinusitis among U.S. children annually. Non-PCV13 serotypes in PCV20 (8, 10A, 11A, 12F, 15B, 22F, and 33F) account for an estimated 1,098,000 outpatient visits and 968,000 antibiotic prescriptions annually. ARIs attributable to PCV15- and PCV20-additional serotypes account for 0.4% and 2.1%, respectively, of all antibiotic prescribing in children. Sensitivity analyses restricted to studies undertaken ≥4 years after PCV13 implementation indicate these primary results may underestimate the burden of ARIs associated with PCV15/20-additional serotypes due to increases in their prevalence through serotype replacement in outpatient disease, although post-PCV13 serotype replacement has not driven considerable increases in U.S. IPD cases.^36^ Among conditions, AOM accounted for the largest burden of all-cause visits and antibiotic prescriptions, consistent with previous studies,^4,5^ and consequently the greatest number of visits and antibiotic prescriptions attributable to PCV15/20-additional serotypes.

In these analyses, PCV20-additional serotypes accounted for at least five times the incidence of outpatient visits and antibiotic prescriptions compared with PCV15-additional serotypes. These findings present a notable contrast to IPD surveillance data, where IPD due to 22F and 33F, found in both PCV15 and PCV20, exceeds the burden of all serotypes in PCV20 not in PCV15.^19^ The pattern in outpatient ARIs, in part, reflects the predominance of 15B/C and 11A carriage among children with ARIs. Among studies included in our analyses, serotype 15B/C alone accounted for 6% of pneumococcal isolates in nasopharyngeal carriage among children experiencing ARIs. A recent review of pneumococcal serotype distribution in MEF and nasopharyngeal carriage in pediatric AOM across countries identified 15B/C as a dominant serotype in 13 out of 17 study populations.^15^ Our findings indicate that PCV15 and PCV20 may offer differential coverage of serotypes contributing to pediatric ARIs, in contrast with IPD.

Because differences in design, enrollment criteria, and microbiologic methods across studies of ARI etiology could preclude generalization of findings to all outpatient ARI cases, our primary analyses used a vaccine-probe design. Attributable fraction estimates based on this approach for AOM were similar to those estimated in analyses based on data from MEF samples or differential carriage of pneumococcal serotypes (children with AOM versus healthy children). For pneumonia and sinusitis we observed greater variability in estimates generated by different methods, with attributable fraction estimates of 0.4–1.6% for PCV15-additional serotypes and 2.7–10.4% for PCV20-additional serotypes. This wide range reflects uncertainty from limited availability of ARI etiology studies in children. However, visit and antibiotic prescription incidence rates for these conditions were low compared with AOM, especially in young children; impacts of PCV15 and PCV20 on pediatric outpatient healthcare utilization will likely be driven by AOM.

Our study has limitations. First, we used ICD-10-CM codes to identify ARIs and could not validate diagnosis accuracy. Second, we used meta-analyses of multiple studies, including studies from non-U.S. high-income countries with similar histories of PCV13 implementation, to estimate serotype distribution and pneumococcal prevalence. Although these measures may vary across settings, the use of multiple studies mitigated bias from single studies or geographic regions. Studies of pediatric ARI etiology in representative samples of U.S. children are needed to monitor pneumococcal disease and inform vaccine formulations. Third, our analyses included estimates of aggregated serotype prevalence in primary studies, although sensitivity analyses showed resulting bias to be low. Cross-protection with PCVs has been observed within certain serogroups (e.g., 6A/6C), although not all (e.g., 19A/19F).^37^ Notably, antibody cross-reaction has been observed for 15B/C, an important contributor to ARIs in our study, but not 15A.^38^ Fourth, although we use negative-controls to account for trends unrelated to vaccination, secular factors not captured by these controls could cause residual bias, especially for sinusitis, for which new diagnostic criteria were introduced in 2014.^26^ Fifth, our analyses address the burden of disease associated with PCV15- and PCV20-additional serotypes, and do not quantify the incidence of disease preventable by vaccination.^24^ While trials have demonstrated that PCV15/20 are safe and immunogenic in children, VE against ARIs is unknown. Vaccine impact will depend on direct and indirect protection and vaccine coverage. Sixth, we do not consider the additional serotypes in PPSV23, which is currently only recommended for select children and is not known to prevent carriage and induce herd protection.^39^ Seventh, long-term PCV use among U.S. children presents challenges in determining total burdens from PCV15/20 serotypes. While replacing PCV13 with PCV10 was associated with increased serotype 3 and 19A disease in Belgium,^35^ dynamics of serotypes under scenarios without PCV use in the United States is unknown. Finally, our study uses data from before the COVID-19 pandemic. Long-term changes in outpatient healthcare utilization and antibiotic prescribing following acute phases of the pandemic are unknown.

## Conclusion

Additional serotypes in PCV15 and PCV20 are estimated to account for 197,000 and 1,098,000 outpatient visits and 173,000 and 968,000 outpatient antibiotic prescriptions, respectively, for AOM, pneumonia, and sinusitis among U.S. children annually, with the greatest burdens among children aged <5 years. Compared with PCV15-additional serotypes, we estimated PCV20-additional serotypes account for >5 times the burden of visits and antibiotic prescriptions. Our findings demonstrate a basis for PCV15 and PCV20, especially PCV20, in preventing ARIs and averting antibiotic use in children.

## Acknowledgements

The authors would like to thank the members of the ACIP Pneumococcal Vaccines Workgroup for their contributions in informing the vaccine-probe approach used in this study. The authors would additionally like to thank Emma Grace Turner for her assistance in literature reviews related to pneumococcal detection and serotype distribution.

## Author contributions

Ms. King had full access to all the data in the study and takes responsibility for the integrity of the data and the accuracy of the data analysis.

Concept and design: King, Lewnard, Andrejko, Kobayashi, Tartof.

Acquisition, analysis, or interpretation of data: King, Lewnard.

Drafting of the manuscript: King, Lewnard, Andrejko, Kobayashi.

Critical revision of the manuscript for important intellectual content: Cohen, Hicks, Kabbani, Tartof.

Statistical analysis: King, Lewnard.

Obtained funding: All authors.

Administrative, technical, or material support: Kabbani, Hicks, Andrejko, Kobayashi.

Supervision: Lewnard, Kobayashi, Cohen, Kabbani, Tartof.

## Disclaimer

The findings and conclusions in this report are those of the authors and do not represent the official position of the Centers for Disease Control and Prevention.

## Previous presentation of reported information

A preliminary version of this analysis using data from 2016– 2018 was presented at the February 2023 Advisory Committee on Immunization Practices Meeting.

## Data availability

MarketScan data is proprietary and therefore data from this study cannot be shared. NAMCS/NHAMCS data are available for public download at https://www.cdc.gov/nchs/ahcd/datasets_documentation_related.htm.

## Use of artificial intelligence and large language models

Artificial intelligence and large language models were not used to create content or assist with writing or editing of this manuscript.

