## Supplemental materials for "Pediatric outpatient visits and antibiotic use attributable to higher valency pneumococcal conjugate vaccine serotypes"

**eMethods 1.** Incidence of outpatient visits and antibiotic prescriptions among children for acute otitis media, pneumonia, and sinusitis due to all causes

**eMethods 2.** Vaccine probe analysis for attributable fractions in AOM

**eMethods 3.** Vaccine probe analysis for non-AOM ARIs

**eMethods 4.** Meta-analysis of pneumococcal serotype distribution

**eMethods 5.** Supplemental approaches for attributable fractions

**eMethods 6.** Sensitivity analysis – serotype distribution ≥4 years post-PCV13 implementation

**eTable 1.** Study data sources and included outpatient settings

**eTable 2.** All cause visits and antibiotic prescriptions from outpatient settings excluding physician offices and emergency departments, MarketScan Databases, 2016–2019

**eTable 3.** *International Classification of Diseases, Tenth Revision, Clinical Modification* codes for conditions of interest

**eTable 4.** Reductions in negative control conditions and all-cause disease pre- and post-PCV13 implementation,[a] vaccine probe analyses

**eTable 5.** Studies included in meta-analysis of pneumococcal serotype distribution in nasopharyngeal samples from children with ARI

**eTable 6.** Serotypes contained in pneumococcal conjugate vaccine inclusion categories

**eTable 7.** Studies included in meta-analysis of pneumococcal detection in nasopharyngeal samples from healthy children and children with ARI

**eTable 8.** Studies included in meta-analyses of pneumococcal detection and serotype distribution in middle ear fluid from children with AOM

**eTable 9.** Serotype distribution by PCV inclusion category and source among samples with pneumococcal detection

**eTable 10.** Proportion of outpatient ARIs attributable to all pneumococcal, PCV15-additional, and PCV20-additional serotypes by condition and estimation method

**eTable 11.** National projection of outpatient visits attributable to any pneumococcal, PCV15-additional, and PCV20-additional serotypes, 2016–2019

**eTable 12.** National projection of antibiotic prescriptions attributable to any pneumococcal, PCV15-additional, and PCV20-additional serotypes, 2016–2019

**eTable 13.** Estimated incidence and annual number of all antibiotic prescriptions among U.S. children, 2016-2019

**eTable 14.** Sensitivity analysis of vaccine serotype distribution ≥4 years and ≥6 years after PCV13 implementation – serotype distribution among samples with pneumococcal detection

**eTable 15.** Sensitivity analysis of serotype distribution ≥4 years after PCV13 implementation – proportion of outpatient ARIs attributable to PCV15- and PCV20-additional serotypes

**eTable 16.** Serotype categorization sensitivity analysis – serotype distribution among samples with pneumococcal detection excluding aggregated serotypes and serogroup counts

**eTable 17.** Serotype categorization sensitivity analysis – proportion of ARIs attributable to PCV15- and PCV20-additional serotypes excluding aggregated serotype and serogroup counts

**eTable 18.** Proportion of ARIs attributable to any serotype in PCV15 and PCV20, including serotypes contained in PCV13

**eTable 19.** Estimated incidence of outpatient visits and antibiotic prescriptions attributable to all serotypes in PCV15 and PCV20, 2016–2019

**References for online only material**

### eMethods 1. Incidence of outpatient visits and antibiotic prescriptions among children for acute otitis media, pneumonia, and sinusitis due to all causes

*eMethods 1.1: Incidence in physician offices and emergency departments.*

We used National Ambulatory Medical Care Survey and National Hospital Ambulatory Medical Care Survey (NAMCS/NHAMCS) data to estimate rates of visits and antibiotic prescribing in physician offices and emergency departments for all U.S. children. These datasets are generated through multi-stage, nationally-representative surveys of healthcare providers administered by the U.S. Centers for Disease Control and Prevention National Center for Healthcare Statistics, and encompass data on medications (≤30 per visit), diagnosis codes (≤5 per visit), and medical services provided for randomly-sampled visits to non-federally employed physician offices and emergency departments (EDs). We used survey-provided sampling weights to generate national estimates of the total numbers of visits to, and antibiotic prescriptions from, physician offices and ED settings from sampled visits. We used data from 2016 and 2019; 2017 data were not available in NAMCS and data integrity concerns precluded use of the 2018 NAMCS nationally-projected estimates.[1] Multiple years of data were required to ensure sufficient sample size for valid national projections. To propagate uncertainty, we defined the estimated national numbers of visits and antibiotic prescriptions within each analysis stratum (age stratum and condition) as Gamma-distributed random variables with variance defined as the square of the sampling error.

*eMethods 1.2: Incidence in all other outpatient settings.*

NAMCS/NHAMCS are limited to physician office and ED settings. Data from the MarketScan Commercial and Medicaid Databases enabled us to further account for healthcare delivery in other outpatient settings (e.g., urgent care, retail health clinics; detailed in **eTable 1**). The MarketScan Databases contain reconciled claims from a convenience sample of >22 million individuals covered by employer-group health insurance and Medicaid-insured individuals in 10 unidentified U.S. states. We considered all claims from a patient on a single day to be part of one visit. To avoid double-counting encounters and prescriptions, we excluded visits to nontraditional outpatient settings occurring on the same day as a physician office or emergency department visit, as we could not reliably determine from which setting an antibiotic prescription originated. We assigned each antibiotic prescription to the most recent outpatient visit within a four-day window (day of prescription and 3 days prior). Members were eligible to be included in the visit/prescription numerator and enrollee denominator if they had continuous medical and prescription coverage in MarketScan for the date of the outpatient visit and subsequent 3 days. We excluded visits from settings from which antibiotic prescriptions are unlikely to occur: independent laboratories, pharmacies, mass immunization centers, and ambulances. We defined counts of outpatient visits and antibiotic prescriptions arising from other outpatient settings (not physician offices or EDs) as Poisson-distributed random variables parameterized by rates observed within these data. We estimated visit and antibiotic prescription incidence rates from MarketScan from 2016–2019.

The MarketScan Commercial and Medicaid Databases represent healthcare encounters for individuals with commercial and public insurance. To estimate national rates of visits and antibiotic prescriptions incidence in nontraditional outpatient settings across these insurance types, we took a weighted average of incidence estimates for each outcome among children with and without commercial insurance, as observed in the MarketScan Commercial and Medicaid Databases (**eTable 2**). Weights were taken from national estimates of the proportion of the pediatric population covered by commercial insurance in 2018 (61.8% of U.S. children 0–19 were commercially insured).[2]

*eMethods 1.3: Total all-cause outpatient incidence estimation.*

To obtain total incidence rates overall and for each age group for both visits and antibiotic prescriptions, we standardized all incidence estimates per 1000 population and summed the rates from office and emergency departments (from NAMCS/NHAMCS) and the weighted averages of rates from all other outpatient settings (from MarketScan). Visits and prescriptions associated with AOM, pneumonia, and sinusitis were identified using *International Classification of Diseases, Tenth Revision, Clinical Modification* (ICD-10-CM) diagnosis codes using an algorithm that assigns a single diagnosis to each visit based on the diagnosis code most likely to result in an antibiotic prescription.[3–5] Briefly, diagnoses were assigned based on the first-listed, lowest-tier diagnosis code, with tiers categorized as: Tier 1 – Antibiotics almost always indicated (miscellaneous bacterial infections, pneumonia, urinary tract infection); Tier 2 – Antibiotics sometimes indicated (AOM, sinusitis, pharyngitis, acne, skin/cutaneous/mucosal infections, gastrointestinal infections, genitourinary infections, acute exacerbations of chronic bronchitis); and Tier 3 – Antibiotics rarely indicated (bronchitis/bronchiolitis, viral upper respiratory infection, nonsuppurative otitis media, influenza, asthma/allergy, fever, other). ICD-10-CM codes for conditions included in this study are in **eTable 3**.

### eMethods 2. Vaccine probe analysis for attributable fractions in AOM

*eMethods 2.1: Attributable fraction for 13-valent pneumococcal conjugate vaccine (PCV13) serotype pneumococci.*

In vaccine-probe studies, the proportion of disease attributable to a vaccine-targeted pathogen can be estimated by dividing vaccine effectiveness (VE) against all-cause disease by VE against the vaccine-targeted pathogen.[6] Thus, studies estimating VE against both outcomes are necessary. No such PCV13 studies with data from children in the United States are available. Therefore, we used data from a long-term surveillance study of AOM incidence and etiology from 2004 to 2015 from southern Israel[7] to define VE against both PCV13 serotype and all-cause AOM. Similar to the United States, PCV13 replaced PCV7 in Israel in 2010 with vaccine coverage among Israeli infants equal to or better than coverage among U.S. infants.[8,9] In this study, middle ear fluid (MEF) samples were taken from children <3 years of age presenting with AOM to the region's only major medical center. Samples were collected from tympanocentesis or spontaneous drainage for culture, and cases were categorized by etiology: *Streptococcus pneumoniae*, non-typeable *Haemophilus influenzae*, *Moraxella catarrhalis*, *Streptococcus pyogenes*, or culture-negative infections. Cases attributable to pneumococci were further categorized by serotype: PCV7 serotypes + 6A, PCV13 serotypes (excluding 6A) not included in PCV7, and non-PCV13 serotypes.

We defined the incidence rate ratio of AOM attributable to PCV13-serotype pneumococci ($\theta_{S13}$) as

$$\theta_{S13} = \frac{\lambda_{S13}(1)}{\lambda_{S13}(0)} \quad , \qquad (1)$$

where $\lambda_{S13}(1)$ represents the observed incidence rate of PCV13-serotype AOM in the presence of a mature PCV13 implementation program (July 2013–June 2015) and $\lambda_{S13}(0)$ represents the observed incidence rate of PCV13-serotype AOM before PCV13 implementation (July 2009–June 2011).

Our vaccine-probe approach to estimating the fraction of AOM attributable to PCV13 serotypes aimed to account for potential secular changes unrelated to PCV13 implementation in AOM incidence over the study period, during which reductions were observed in incidence of AOM due PCV13-serotype pneumococci as well as cases due to other etiologies (**eTable 4**). To account for secular trends that may have also contributed to declining AOM incidence during this time period[10] we further measured the incidence rate ratio of AOM attributable to all other etiologies ($\theta_N$) as

$$\theta_N = \frac{\lambda_N(1)}{\lambda_N(0)} \quad , \qquad (2)$$

where $\lambda_N(1)$ and $\lambda_N(0)$ represent observed incidence rates of AOM due to all non-pneumococcal etiologies during July 2013–June 2015 and July 2009–June 2011, respectively. We did not include non-vaccine type pneumococcal AOM in our negative control group due to the potential for vaccine-driven serotype replacement to contribute to changes in incidence of AOM due to these serotypes. To exclude the secular change from estimates of the reduction in PCV13-serotype disease attributable to implementation of PCV13,[11] we divided the incidence rate ratio of PCV13 serotype-attributable AOM ($\theta_{S13}$) by the incidence rate ratio of AOM attributable to all other etiologies ($\theta_N$) and defined VE against PCV13-serotype disease ($\mathrm{VE}_{S13}$) as

$$\mathrm{VE}_{S13} = 1 - \frac{\theta_{S13}}{\theta_N} \quad . \qquad (3)$$

We also accounted for changes in incidence driven by secular factors when estimating the effect of PCV13 on incidence of AOM due to all causes. Defining $\lambda_{AC}(1)$ and $\lambda_{AC}(0)$ as the observed incidence rates for all-cause AOM during the periods of July 2013–June 2015 and July 2009–June 2011 within the study setting, respectively, we estimated $\lambda_{AC}^*(1)$, the counterfactual incidence rate of all-cause AOM incidence during July 2013–June 2015 resulting from a continuation of secular trends, without PCV13 introduction, as

$$\lambda_{AC}^*(1) = \theta_N \lambda_{AC}(0) \quad . \qquad (4)$$

Based on this estimate of the counterfactual incidence of all-cause AOM absent PCV13 implementation, we estimated VE against all-cause AOM ($\mathrm{VE}_{AC}$) as

$$\mathrm{VE}_{AC} = 1 - \frac{\lambda_{AC}(1)}{\lambda_{AC}^*(1)} \quad , \qquad (5)$$

with the difference $\lambda_{AC}^*(1) - \lambda_{AC}(1)$ corresponding to the absolute incidence of AOM prevented by PCV13 implementation within the study setting. Finally, we estimated the attributable fraction of PCV13-serotype AOM cases, absent PCV13 implementation ($\mathrm{AF}_{13}^*$), as:

$$\mathrm{AF}_{13}^{*} = \frac{\mathrm{VE}_{\mathrm{AC}}}{\mathrm{VE}_{\mathrm{S13}}} \quad , \tag{6}$$

and defined the incidence of PCV13-serotype AOM within the study setting, absent PCV13 implementation, as $\lambda_{13}^{*}(1) = \mathrm{AF}_{13}^{*}\lambda_{AC}^{*}(1)$.

*eMethods 2.2. Attributable fractions for additional serotypes included in PCV15/PCV20 not included in PCV13.*

We used data on serotype distributions of AOM in high-income settings with PCV13 programs to estimate the proportion of disease cases attributable to serotypes targeted by PCV15 and PCV20 (see **eMethods 4** for further details on the literature review and estimation framework). Incorporating these data, we defined $p_{13}$ as the residual proportion of all pneumococcal AOM cases due to PCV13 serotypes such settings, $p_{15}$ as the proportion due to all serotypes included in PCV15 (residual PCV13 serotypes plus 22F and 33F), and $p_{20}$ as the proportion due to all serotypes included in PCV20 (residual PCV13 serotypes plus 8, 10A, 11A, 12F, 15B/C, 22F, and 33F). We defined the proportions attributable to PCV15-additional and PCV20-additional serotypes as $\alpha_{15} = p_{15} - p_{13}$ (for serotypes 22F and 33F) and $\alpha_{20} = p_{20} - p_{13}$ (for serotypes 8, 10A, 11A, 12F, 15B/C, 22F, and 33F), respectively.

Thus, we defined $\lambda_{15+}(1)$ and $\lambda_{20+}(1)$ as the incidence of AOM attributable to the additional serotypes in PCV15 (22F, 33F) and PCV20 (8, 10A, 11A, 12F, 15B/C, 22F, and 33F ), respectively, under present circumstances of PCV13 use as

$$\lambda_{15+}(1) = \frac{\alpha_{15}}{p_{13}}\lambda_{S13}(1) \tag{7a}$$

and

$$\lambda_{20+}(1) = \frac{\alpha_{20}}{p_{13}}\lambda_{S13}(1) \quad . \tag{7b}$$

The above rates, $\lambda_{15+}(1)$ and $\lambda_{20+}(1)$, can be interpreted as the additional disease burden caused by serotypes targeted by PCV15/20 in comparison to PCV13. Thus, the fraction of existing all-cause AOM ($\lambda_{AC}(1)$) attributable to PCV15/20-additional serotypes under present circumstances are

$$\mathrm{AF}_{15+} = \frac{\lambda_{15+}(1)}{\lambda_{AC}(1)} \tag{8a}$$

and

$$\mathrm{AF}_{20+} = \frac{\lambda_{20+}(1)}{\lambda_{AC}(1)} \quad . \tag{8b}$$

*eMethods 2.3. Attributable fractions for all serotypes included in PCV15/PCV20.*

We further aimed to estimate the total incidence of AOM attributable to all PCV15/20-targeted serotypes, including disease prevented by PCV13 and PCV13 residual disease to quantify the total disease burden targeted by PCV15/20. We defined these rates as

$$\lambda_{15}^{*}(1) = \lambda_{13}^{*}(1) + \lambda_{15+}(1) \tag{9a}$$

and

$$\lambda_{20}^{*}(1) = \lambda_{13}^{*}(1) + \lambda_{20+}(1) \quad , \tag{9b}$$

when considering both the burden of disease expected to result from PCV13 serotypes (prevented by PCV13 and residual), $\lambda_{13}^{*}(1)$, and the observed burden of PCV15/20 additional serotypes under present circumstances, $\lambda_{15+}(1)$ and $\lambda_{20+}(1)$. Thus, the corresponding attributable fractions are

$$\mathrm{AF}_{15}^{*} = \frac{\lambda_{15}^{*}(1)}{\lambda_{AC}^{*}(1)} \tag{10a}$$

and

$$\mathrm{AF}_{20}^{*} = \frac{\lambda_{20}^{*}(1)}{\lambda_{AC}^{*}(1)} \quad . \tag{10b}$$

To estimate AOM incidence due to all PCV15 serotypes and PCV20 serotypes, we multiply observed rates of AOM incidence due to all causes by the terms $\frac{\lambda^*_{AC}(1)}{\lambda_{AC}(1)}$ and $\mathrm{AF}^*_{15}$ or $\mathrm{AF}^*_{20}$, respectively, to account for both the expected relative rate of incidence of AOM due to all causes, including all PCV13 disease ($\lambda^*_{AC}(1)/\lambda_{AC}(1)$) and the fraction of cases expected to be attributable to all PCV15/20 serotypes ($\mathrm{AF}^*_{15}$ and $\mathrm{AF}^*_{20}$).

### eMethods 3. Vaccine probe analysis for non-AOM ARIs

We used a framework similar to our vaccine probe analyses of AOM (**eMethods 2**) to determine PCV15/20-additional and total serotype etiologic fractions for other (non-AOM) ARIs. To estimate PCV13 VE against all-cause pneumonia and sinusitis visits, we compared incidence rates before (2009–10) and after (2013–15) PCV13 implementation in the United States among children aged ≤3 years using data from NAMCS/NHAMCS. We selected these time periods and age groups to mirror our analyses of PCV13 VE in AOM.[7] Because no eligible published studies of sinusitis and outpatient pneumonia etiologies in children were available in the post-PCV13 period, we assumed that PCV13 VE against outpatient pneumonia and sinusitis attributable to PCV13-targeted serotypes was equivalent to that observed for AOM.

We used skin and soft tissue infections (SSTI) as a negative-control condition to account for secular trends in visit incidence unrelated to PCV13 (e.g., changes in healthcare seeking behavior; **eTable 4**). We divided incidence rate ratios for all-cause pneumonia and sinusitis visits in 2013–15 vs. 2009–10 by incidence rate ratios for SSTI visits in 2013–15 vs. 2009–10 to adjust estimates of PCV13 impact for secular trends.[11] We then estimated PCV13 and PCV15/20 attributable proportions for pneumonia and sinusitis using the same framework employed for AOM (**equations 7–10**), estimating the incidence of pneumonia and sinusitis due to PCV13 serotypes, in the context of a mature PCV13 program, as

$$\lambda_{13}(1) = \mathrm{AF}^{*}_{13}\theta_N\lambda_{AC}(0) - [\theta_N\lambda_{AC}(0) - \lambda_{AC}(1)] \ ,$$

where AF $*_{13}$ $\theta_N\lambda_{AC}(0)$ is the estimated incidence of PCV13 serotype disease had PCV13 not been implemented and $\theta_N\lambda_{AC}(0) - \lambda_{AC}(1)$ is the amount of PCV13-prevented disease. Consistent with the framework outlined in **eMethods 2**, $\theta_N$ denotes the incidence rate ratio of the negative control condition (here, SSTI) comparing incidence in 2013–15 to 2009–10. Vaccine serotype category proportions were estimated using serotype data from nasopharyngeal samples in children experiencing AOM and other ARIs as detailed in **eMethods 4**; the majority of studies used to estimate serotype proportions were among children with AOM (**eTable 5**).

### eMethods 4. Meta-analysis of pneumococcal serotype distribution

We obtained estimates of pneumococcal serotype distributions in nasopharyngeal carriage among children with AOM and other ARIs via meta-analyses of existing studies. We included studies meeting the following criteria: 1) pediatric study population, 2) study period ≥1 year after PCV13 implementation, 3) study population from high-income countries where only PCV13 was recommended for infants (rather than exclusive PCV10 or mixed PCV10/13 programs). We included studies undertaken outside the U.S. due to limited numbers and geographic scope of U.S.-based studies. Consistency in estimates of pneumococcal prevalence and serotype distribution in both historic[12,13] and present studies[14–37] of ARI supported this approach. Where multiple studies included overlapping study populations, we included only non-overlapping strata, or included the study with the most comprehensive study population. Only one study estimated serotype distribution in non-AOM ARI (lower respiratory tract infection).[16] Therefore, we included AOM studies for our estimates of serotype distribution in non-AOM ARIs. The included studies are presented in **eTable 5.**

We grouped serotypes by non-overlapping PCV inclusion categories (PCV13, PCV15-additional [22F, 33F], PCV20, non-PCV15-additional [8, 10A, 11A, 12F, 15B/C], and non-vaccine type) and extracted isolate counts by vaccine category from the published studies identified in our literature review. We defined the proportion of isolates belonging to each category as a Dirichlet distribution, for which we sampled parameters via Markov chain Monte Carlo (MCMC) sampling using data on isolate counts from each study. We fit Dirichlet distributions separately for serotypes represented in 1) nasopharyngeal carriage in children with AOM, and 2) nasopharyngeal carriage in children with AOM and other ARIs. Mutually exclusive PCV inclusion categories were required for this MCMC approach; we then summed estimates from the PCV20, non-PCV15-additional and PCV15-additional groups to estimate the proportion of isolates represented by PCV20-additional serotypes (8, 10A, 11A, 12F, 15B/C, 22F, 33F). Few studies were available for children >5 years of age; we assumed serotype distribution was consistent across age groups.

Published studies included in our meta-analysis of serotype distributions presented serotype data with heterogeneous granularity. For studies presenting data on serotypes individually, we included only vaccine-type serotypes in our categorization; however, when studies provided aggregated serogroup or serotype counts for serotypes grouped across multiple relevant categories, we included the aggregated counts in the relevant PCV inclusion category. For example, if a study presented combined counts for 22A/F we included the aggregated count in the PCV15-additional serotype category. However, if a study presented counts separately for 22A and 22F, we included only the 22F counts in the PCV15 category. The categorization scheme is presented in **eTable 6**. While this approach is supported by evidence for intra-serogroup cross-protection for certain serotypes,[38,39] it may nonetheless overestimate vaccine coverage across serotypes. We therefore conducted a sensitivity analysis using only isolate counts from strictly defined vaccine serotype groups and counting multi-serotype aggregated strata from the included studies as non-vaccine serotypes only. Results of this sensitivity analysis are presented in **eTables 16-17**.

### eMethods 5. Supplemental approaches for attributable fractions

*eMethods 5.1. PCV15/20-additional serotype attributable fractions in AOM based on data from middle ear fluid (MEF).*

We first established the proportion of all cases of AOM attributable to any pneumococci via meta-analysis of published estimates of pneumococcal prevalence in MEF samples from children with AOM, conducted using the Metafor package for R.[40] We then conducted a meta-analysis analogous to that described in **eMethods 4** focused on studies with serotype data from studies sampling MEF in children with AOM to establish the corresponding serotype distribution. Study inclusion criteria for both MEF pneumococcal prevalence and serotype distribution meta-analyses were consistent with the main analysis serotype distribution meta-analysis, as described in **eMethods 4**. Included studies for both pneumococcal prevalence and serotype distribution are presented in **eTable 8**. Finally, to obtain the fraction of cases attributable to PCV15/20-additional serotypes, we multiplied the proportion of cases attributable to pneumococci by the proportion of all pneumococcal isolates accounted for by each PCV inclusion category.

*eMethods 5.2. PCV15/20-additional serotype attributable fractions in AOM estimated via a differential carriage approach.*

We similarly obtained estimates of pneumococcal prevalence in nasopharyngeal carriage among children with AOM and other ARIs and in healthy children using meta-analyses of existing studies. Healthy children were defined as those presenting for well-visits, presenting for injuries or illnesses unrelated to ARI, and those sampled based on presence in daycare. We included AOM studies to generate estimates of pneumococcal prevalence in non-AOM ARIs as only one study estimated pneumococcal prevalence in pneumonia[23] and none examined sinusitis. The included studies are presented in **eTable 7.** We used meta-regression to estimate the difference in pneumococcal carriage prevalence between healthy children and those experiencing either AOM or other ARIs, which we defined as the fraction of AOM attributable to (any) pneumococci. We defined meta-regression models adjusted for years since PCV13 implementation, country, and laboratory method used to identify pneumococcus (phenotypic versus molecular assays) as covariates. We conducted all meta-regression analyses using the Metafor package for R.[40] We multiplied pneumococcal attributable fractions established via meta-regression by the proportion of all pneumococcal isolates accounted for by each PCV inclusion category from **eMethods 4** (included studies available in **eTable 5**) to establish PCV15- and PCV20-additional serotype attributable fractions.

### eMethods 6. Sensitivity analysis – serotype distribution ≥4 years post-PCV13 implementation

Our meta-analysis included serotype distribution data from studies beginning one year post-PCV13 implementation in order to include the most comprehensive scope of studies. This approach likely yielded conservative estimates of PCV15/20-additional serotype proportions given that decreases in PCV13 serotypes and serotype replacement after PCV13 increased the representation of PCV15/20-additional serotypes over time. In children, serotype distribution stabilization has been generally been noted within four to six years after PCV implementation, although greater variation has been observed in adult disease.[41–43] We therefore conducted a sensitivity analysis including only published estimates from study periods beginning ≥4 years after PCV13 implementation. Results using data from this sensitivity analysis are presented in **eTables 14–15**.

### eTable 1. Study data sources and included outpatient settings

| Data source | Outpatient care delivery setting(s) included in study |
|---|---|
| National Ambulatory Care Medical Survey (NAMCS) | Physician office |
| National Hospital Ambulatory Care Medical Survey (NHAMCS) | Emergency department |
| MarketScan Commercial and Medicaid Databases[a] | Outpatient hospital facility, urgent care facility, retail health clinic, telehealth, independent clinic, Tribal 638 facility, Indian Health Services facility, rural health clinic, public health clinic, federally-qualified health center, military treatment facility, school, inpatient hospital, ambulatory surgery center, birthing center, place of employment, patient home/lodging, prison, homeless shelter, mobile unit service, end stage renal disease facility, rehabilitation facility, assisted living, long-term care/nursing facility, skilled nursing facility, other residential facility,[b] hospice, psychiatric facility, community mental health center, other/unknown outpatient facility |
| Excluded | Pharmacy, independent laboratory, mass immunization center, ambulance |

[a] For care delivery settings providing both inpatient and outpatient care, only outpatient services included.
[b] Other residential facility includes custodial care facility, adult living facility, residential substance abuse facility, group home, intellectual disability care facility.

**eTable 2. All-cause visits and antibiotic prescriptions from outpatient settings excluding physician offices and emergency departments,[a] MarketScan Databases, 2016–2019**

| | | Incidence per 1000 person years (95% CI) | | | |
|---|---|---|---|---|---|
| | | *Outpatient visits* | | *Antibiotic prescriptions* | |
| **Age stratum** | **Condition** | Commercially insured children | Non-commercially insured children | Commercially insured children | Non-commercially insured children |
| <2 years | | | | | |
| | AOM | 76 (60, 94) | 113 (93, 134) | 43 (31, 56) | 77 (60, 95) |
| | Pneumonia | 6 (2, 12) | 9 (4, 16) | 2 (0, 6) | 3 (0, 6) |
| | Total[b,c] | 83 (65, 101) | 122 (101, 144) | 45 (33, 59) | 80 (63, 98) |
| 2–4 years | | | | | |
| | AOM | 45 (33, 59) | 79 (62, 97) | 31 (21, 42) | 53 (39, 68) |
| | Pneumonia | 6 (2, 11) | 7 (3, 13) | 2 (0, 6) | 3 (0, 7) |
| | Total[b,c] | 51 (38, 66) | 86 (69, 105) | 34 (23, 46) | 56 (42, 72) |
| All children <5 years | | | | | |
| | AOM | 57 (43, 73) | 91 (72, 110) | 36 (24, 48) | 61 (46, 77) |
| | Pneumonia | 6 (2, 11) | 8 (3, 14) | 2 (0, 6) | 3 (0, 7) |
| | Total[b,c] | 63 (48, 80) | 98 (80, 119) | 38 (27, 51) | 64 (49, 80) |
| 5–17 years | | | | | |
| | AOM | 11 (5, 18) | 18 (11, 28) | 9 (3, 15) | 14 (7, 22) |
| | Pneumonia | 2 (0, 6) | 3 (0, 7) | 1 (0, 4) | 1 (0, 4) |
| | Sinusitis | 8 (3, 14) | 10 (5, 17) | 6 (2, 11) | 8 (3, 14) |
| | Total[b,c] | 22 (13, 31) | 32 (22, 44) | 16 (9, 24) | 23 (15, 34) |
| All children ≤17 years | | | | | |
| | AOM | 22 (13, 32) | 38 (26, 51) | 15 (8, 23) | 27 (17, 37) |
| | Pneumonia | 3 (0, 8) | 4 (1, 9) | 5 (1, 10) | 2 (0, 5) |
| | Sinusitis | 7 (3, 13) | 10 (4, 17) | 1 (0, 4) | 7 (3, 13) |
| | Total[b,c] | 33 (22, 44) | 53 (39, 67) | 22 (13, 32) | 36 (25, 49) |

Abbreviations: CI – confidence interval; AOM – acute otitis media

[a] Settings include retail health clinics, urgent care facilities, outpatient hospital departments, telehealth, and other. All outpatient settings detailed in **eTable 1**.

[b] Incidence by condition may not sum to total due to rounding.

[c] Estimates for sinusitis not included in age-stratum totals for <2 years, 2–4 years, and all <5 years as sampled visit counts for sinusitis do not meet National Ambulatory Medical Care Survey and National Hospital Ambulatory Medical Care Survey minimum sample size requirements for valid projection to national estimates.[1] Sinusitis is uncommon in children <5 years due to sinus development.[44,45] To generate total estimates for all conditions across all children, visits, and antibiotic prescriptions for sinusitis in children <5 years were included in the sinusitis and total estimates.

### eTable 3. *International Classification of Diseases, Tenth Revision, Clinical Modification* (ICD-10-CM) codes for conditions of interest[a]

| Condition | ICD-10-CM diagnosis codes |
|---|---|
| Acute otitis media | H66*, H67*, H73.0*, H73.1*, H73.2* H73 |
| Pneumonia | A48.1, B01.2, B05.2, B06.81,[b] J09.X1,[b] J10.0*, J11.0*, J12*–J18*, J69, J690, J84.116,[b] J95.851,[b] O29.01*,[b] O74.0, O89.01,[b] P23* |
| Sinusitis | J01*, J32* |

* Includes all ICD-10-CM codes under listed parent code.

[a] Diagnosis codes used to identify visits for conditions of interest using tiered diagnosis algorithm that assigns a single condition to a visit based on the most likely antibiotic indication. For complete description of this algorithm and all conditions and associated ICD-10-CM codes, see King et al. 2021.[5]

[b] Only included in MarketScan incidence estimates due to truncation of ICD-10-CM codes in NAMCS/NHAMCS.

### eTable 4. Reductions in negative control conditions and all-cause disease pre- and post-PCV13 implementation,[a] vaccine probe analyses

| Condition | Observed reductions in disease, % (95% CI) | | | Adjusted reduction in all-cause disease, % (95% CI) |
|---|---|---|---|---|
| | All-cause disease | Negative control disease[b] | PCV13 serotype disease[c] | |
| AOM[d] | 62.0 (58.4, 65.4) | 56.1 (51.3, 60.5) | 90.3 (85.8, 93.4) | 13.5 (3.9, 22.1) |
| Pneumonia[e] | 37.9 (37.7, 38.0) | 23.9 (23.8, 24.1) | -- | 18.3 (18.2, 18.4) |
| Sinusitis[e] | 44.8 (44.6, 44.9) | 23.9 (23.8, 24.1) | -- | 27.4 (27.3, 27.5) |

Abbreviations: PCV – pneumococcal conjugate vaccine; CI – confidence interval; AOM – acute otitis media

[a] For AOM, July 2009–June 2011 was considered the pre-PCV13 period and July 2013–June 2015 was considered the post-PCV13 period. For pneumonia and sinusitis, the pre- and post-PCV13 periods were January 2009–December 2010 and January 2013–December 2015, respectively. See **eMethods 2**, **eMethods 3**.

[b] Negative control for AOM is AOM due to non-pneumococcal etiologies. Negative control for pneumonia and sinusitis is skin and soft tissue infections.

[c] Microbiological data comparing pre- and post-PCV13 period only available for AOM. PCV13 serotype reductions assumed to be constant across conditions for vaccine-probe analyses.

[d] Based on data from Ben-Shimol et al. 2016[7] as detailed in **eMethods 2**.

[e] Based on data estimated using the National Ambulatory Medical Care Survey and National Hospital Ambulatory Medical Care Survey for children <3 years as detailed in **eMethods 3.**

### eTable 5. Studies included in meta-analysis of pneumococcal serotype distribution in nasopharyngeal samples from children with ARI

| Reference | Study years | Study country | Participant ages | Condition |
|---|---|---|---|---|
| Alleman 2017[15] | 2011–2015 | Switzerland | All children[a] | AOM |
| Ben-Shimol 2021[16,b] | 2011–2017 | Israel | <2 years | LRTI |
| Chen 2020[17] | 2016–2018 | Taiwan | 2–60 months | AOM |
| Kaur 2022[14] | 2015–2019 | United States | 6–36 months | AOM |
| Martin 2018[27] | 2012–2014 | United States | 6–23 months | AOM |

Abbreviations: ARI – acute respiratory infection; AOM – acute otitis media; LRTI – lower respiratory tract infection

[a] Also includes limited number of adults.

[b] Study also includes AOM sampled from middle ear fluid, which was not considered in the main analysis. Middle ear fluid serotype distribution only used for supplementary analyses. See **eMethods 5**, **eTable 8**.

### eTable 6. Serotypes contained in pneumococcal conjugate vaccine inclusion categories

| Vaccine category | Vaccine serotypes | Related serotypes/ serogroups[a] |
|---|---|---|
| **PCV13** | 1, 3, 4, 5, 6A/B, 7F, 9V, 14, 18C, 19A/F, 23F | |
| **PCV15-additional** | 22F, 33F | 22A/F, 22, 33F/37, 33 |
| **PCV20-additional** | 22F, 33F, 8, 10A, 11A, 12F, 15B/C | 22A/F, 22, 33F/37, 33, 10, 11A/D, 12A/B/F, 15 |

Abbreviations: PCV – pneumococcal conjugate vaccine

[a] Aggregated serotype or serogroup-level isolate counts were included only when isolate counts for individual serotypes were not provided in published studies included in meta-analyses of serotype distribution.

**eTable 7. Studies included in meta-analysis of pneumococcal detection in nasopharyngeal samples from healthy children and children with ARI**

| Reference | Study years | Study country | Participant ages | Disease status[a] | Laboratory method(s) |
|---|---|---|---|---|---|
| Candeias 2022[37] | 2018–2020 | Portugal | <6 years | Healthy | PCR |
| Chen 2020[17] | 2016–2018 | Taiwan | 2–60 months | AOM, Healthy | Culture, PCR |
| Desai 2015[18] | 2010–2013 | United States | 6–59 months | AOM | Culture |
| Dunais 2015[19] | 2012 | France | 3–40 months | Healthy | Culture |
| Kandasamy 2020[20] | 2014–2015 | United Kingdom | 13–48 months | Healthy | Culture |
| Kaur 2022 | 2015–2019 | United States | 6–36 months | AOM, Healthy | Culture |
| Kaur 2016[21] | 2010–2015 | United States | 6–30 months | Healthy | Culture |
| Kovács 2019[35] | 2015–2016 | Hungary | 1–6 years | Healthy | Culture |
| Lewnard 2021[23] | 2009–2018 | Israel | 4–59 months | CAP, Healthy | Culture |
| Lindstrand 2016[24] | 2011–2015 | Sweden | <5 years | Healthy | Culture |
| Løvlie 2020[34] | 2013–2015 | Norway | Children in daycare[b] | Healthy | Culture |
| Mameli 2015[25] | 2011–2012 | Italy | 9–71 months | Healthy | Culture |
| Martin 2018[27] | 2012–2014 | United States | 6–23 months | AOM | Culture |
| Ouldali 2019[28] | 2011–2018 | France | 6–24 months | AOM | Culture |
| Pichichero 2018[30] | 2010–2013 | United States | 6–36 months | AOM | Culture |
| Rybak 2022[31] | 2011–2021 | France | >6 months | Healthy | Culture |
| Tsai 2020[32] | 2013–2017 | Taiwan | 1–36 months | Healthy | Culture |

Abbreviations: ARI – acute respiratory infection; PCR – polymerase chain reaction; AOM – acute otitis media; CAP – community acquired pneumonia

[a] Healthy children were defined as those presenting for well-visits, presenting for injuries or illnesses unrelated to ARI, and those sampled based on presence in daycare.

[b] Study sample included children of any age in daycare.

**eTable 8. Studies included in meta-analyses of pneumococcal detection and serotype distribution in middle ear fluid from children with AOM**

| Reference | Study years | Study country | Participant ages | Pneumococcal prevalence in MEF | Serotype distribution in MEF |
|---|---|---|---|---|---|
| Ben-Shimol 2021[16] | 2011–2017 | Israel | <24 months | | X |
| Grant 2022[36] | 2011–2020 | United States | <5 years | | X |
| Kaur 2022[14] | 2015–2019 | United States | 6–36 months | X | X |
| Levy 2019[22] | 2015–2018 | France | 3 months–15 years | X | X |
| Marchisio 2017[26] | 2015–2016 | Italy | All children | X | X |
| Nakano 2020[29] | 2013–2017 | Japan | 2 months–16 years | | X |
| Pichichero 2018[30] | 2010–2013 | United States | 6–36 months | X | |
| Ubukata 2018[33] | 2016–2017 | Japan | <15 years | X | X |

Abbreviations: AOM – acute otitis media; MEF – middle ear fluid

### eTable 9. Serotype distribution by PCV inclusion category and source among samples with pneumococcal detection[a]

| Sample Source | Percent of all pneumococcal isolates (95% CI) | | |
|---|---|---|---|
| | *PCV13*[b] | *PCV15-additional*[b] | *PCV20-additional*[b] |
| ARI—NP carriage | 23.3 (21.6, 25.1) | 3.6 (2.8, 4.4) | 22.7 (21.0, 24.5) |
| AOM—NP carriage | 24.5 (21.3, 28.0) | 5.2 (3.7, 7.1) | 26.9 (23.5, 30.4) |
| AOM—MEF | 35.5 (33.0, 38.1) | 4.6 (3.6, 5.8) | 22.9 (20.7, 25.1) |

Abbreviations: PCV – pneumococcal conjugate vaccine; CI – confidence interval; ARI – acute respiratory infection; NP – nasopharyngeal; AOM – acute otitis media; MEF – middle ear fluid

[a] Estimated using Markov-chain Monte Carlo approach based on published estimates from studies in **eTable 5**. **Figure 2** represents data by serotype from the same studies (pooled counts).

[b] Includes vaccine serotypes and corresponding aggregated serotypes/serogroups as detailed in **eTable 6**. PCV-15 additional serotypes (not included in PCV13): 22F, 33F. PCV20-additional serotypes: 8, 10A, 11A, 12F, 15B, 22F, 33F.

### eTable 10. Proportion of outpatient ARIs attributable to all pneumococcal, PCV15-additional, and PCV20-additional serotypes[a] by condition and estimation method

| | | Percent of cases attributable to pneumococcal serotypes (95% CI) | | |
|---|---|---|---|---|
| **Condition** | **Estimation method** | *All pneumococci* | *PCV15-additional*[a] | *PCV20-additional*[a] |
| AOM | | | | |
| | Vaccine Probe | 19.4 (17.0, 22.4) | 1.0 (0.7, 1.5) | 5.2 (4.2, 6.5) |
| | Middle Ear Fluid | 22.3 (17.2, 27.4) | 1.0 (0.7, 1.4) | 5.1 (3.9, 6.4) |
| | Differential Carriage | 13.9 (2.0, 25.9) | 0.7 (0.1, 1.5) | 3.7 (0.5, 7.1) |
| Pneumonia | | | | |
| | Vaccine Probe | 27.3 (19.0, 40.3) | 1.0 (0.6, 1.5) | 6.2 (4.2, 9.3) |
| | Differential Carriage[b] | 11.8 (1.0, 22.7) | 0.4 (0.0, 0.8) | 2.7 (0.2, 5.2) |
| Sinusitis | | | | |
| | Vaccine Probe | 45.8 (31.9, 67.5) | 1.6 (1.1, 2.6) | 10.4 (7.1, 15.5) |
| | Differential Carriage[b] | 11.8 (1.0, 22.7) | 0.4 (0.0, 0.8) | 2.7 (0.2, 5.2) |

Abbreviations: PCV – pneumococcal conjugate vaccine; ARI – Acute respiratory infection; CI – confidence interval; AOM – acute otitis media

[a] Includes vaccine serotypes and corresponding aggregated serotypes/serogroups as detailed in **eTable 6**. PCV-15 additional serotypes (not included in PCV13): 22F, 33F. PCV20-additional serotypes: 8, 10A, 11A, 12F, 15B, 22F, 33F.

[b] Consistent for all non-AOM ARIs.

**eTable 11. National projection of outpatient visits attributable to any pneumococcal, PCV15-additional, and PCV20-additional serotypes,[a] 2016–2019**

| | | Estimated annual outpatient visits by U.S. children, in thousands (95% CI) | | |
|---|---|---|---|---|
| **Age stratum** | **Condition** | *All pneumococci* | *PCV15-additional serotypes[a]* | *PCV20-additional serotypes[a]* |
| <2 years | | | | |
| | AOM | 886 (610, 1261) | 46 (27, 76) | 237 (158, 352) |
| | Pneumonia | 94 (49, 173) | 3 (2, 6) | 21 (11, 39) |
| | Total[b] | 983 (701, 1367) | 50 (30, 80) | 260 (178, 376) |
| 2–4 years | | | | |
| | AOM | 818 (571, 1151) | 43 (26, 69) | 220 (147, 322) |
| | Pneumonia | 94 (50, 171) | 3 (2, 6) | 21 (11, 39) |
| | Total[b] | 917 (661, 1259) | 46 (29, 73) | 242 (167, 347) |
| All children <5 years[c] | | | | |
| | AOM | 1715 (1300, 2246) | 90 (57, 138) | 460 (333, 634) |
| | Pneumonia | 190 (113, 319) | 7 (4, 12) | 43 (25, 73) |
| | Total[b] | 1912 (1481, 2461) | 97 (63, 146) | 505 (372, 682) |
| 5–17 years | | | | |
| | AOM | 646 (423, 962) | 34 (19, 57) | 173 (110, 267) |
| | Pneumonia | 147 (77, 268) | 5 (3, 10) | 33 (17, 61) |
| | Sinusitis | 11228 (733, 2052) | 44 (25, 77) | 279 (164, 469) |
| | Total[b] | 2043 (1431, 2971) | 84 (55, 128) | 491 (341, 719) |
| All children ≤17 years[c] | | | | |
| | AOM | 2362 (1794, 3091) | 123 (78, 191) | 634 (458, 872) |
| | Pneumonia | 339 (206, 556) | 12 (7, 21) | 77 (46, 127) |
| | Sinusitis | 1655 (957, 2841) | 59 (32, 106) | 375 (215, 651) |
| | Total[b] | 4392 (3347, 5884) | 197 (133, 287) | 1098 (816, 1489) |

Abbreviations: PCV – pneumococcal conjugate vaccine; CI – confidence interval; AOM – acute otitis media

[a] Includes vaccine serotypes and corresponding aggregated serotypes/serogroups as detailed in **eTable 6**. PCV-15 additional serotypes (not included in PCV13): 22F, 33F. PCV20-additional serotypes: 8, 10A, 11A, 12F, 15B, 22F, 33F.

[b] Number of visits by condition may not sum to total due to rounding.

[c] Number of visits across age groups may not sum to all children totals due to rounding.

### eTable 12. National projection of antibiotic prescriptions attributable to any pneumococcal, PCV15-additional, and PCV20-additional serotypes,[a] 2016–2019

| | | Estimated annual outpatient antibiotic prescriptions among U.S. children, in thousands (95% CI) | | |
|---|---|---|---|---|
| **Age stratum** | **Condition** | *All pneumococci* | *PCV15-additional serotypes[a]* | *PCV20-additional serotypes[a]* |
| <2 years | | | | |
| | AOM | 633 (421, 928) | 33 (19, 55) | 170 (109, 259) |
| | Pneumonia | 71 (43, 118) | 3 (1, 4) | 16 (10, 27) |
| | Total[b] | 706 (490, 1006) | 36 (21, 58) | 186 (124, 277) |
| 2–4 years | | | | |
| | AOM | 747 (535, 1027) | 39 (24, 62) | 200 (138, 288) |
| | Pneumonia | 90 (57, 145) | 3 (2, 5) | 20 (13, 33) |
| | Total[b] | 840 (621, 1125) | 42 (27, 66) | 222 (157, 311) |
| All children <5 years[c] | | | | |
| | AOM | 1387 (1057, 1809) | 72 (46, 111) | 372 (269, 509) |
| | Pneumonia | 162 (103, 259) | 6 (3, 10) | 37 (23, 59) |
| | Total[b] | 1555 (1211, 1991) | 78 (51, 118) | 410 (304, 551) |
| 5–17 years | | | | |
| | AOM | 659 (512, 845) | 34 (22, 52) | 177 (130, 239) |
| | Pneumonia | 122 (78, 194) | 4 (3, 7) | 28 (18, 45) |
| | Sinusitis | 1135 (681, 1885) | 40 (23, 70) | 257 (153, 431) |
| | Total[b] | 1925 (1402, 2744) | 80 (54, 119) | 465 (335, 663) |
| All children ≤17 years[c] | | | | |
| | AOM | 2049 (1606, 2597) | 107 (69, 162) | 550 (408, 737) |
| | Pneumonia | 333 (214, 523) | 12 (7, 20) | 75 (48, 120) |
| | Sinusitis | 1473 (815, 2602) | 52 (28, 97) | 334 (184, 597) |
| | Total[b] | 3881 (2969, 5248) | 173 (118, 252) | 968 (722, 1318) |

Abbreviations: PCV – pneumococcal conjugate vaccine; CI – confidence interval; AOM – acute otitis media

[a] Includes vaccine serotypes and corresponding aggregated serotypes/serogroups as detailed in **eTable 6**. PCV-15 additional serotypes (not included in PCV13): 22F, 33F. PCV20-additional serotypes: 8, 10A, 11A, 12F, 15B, 22F, 33F.

[b] Number of antibiotic prescriptions by condition may not sum to total due to rounding.

[c] Number of antibiotic prescriptions across age groups may not sum to all children totals due to rounding.

**eTable 13. Estimated incidence and annual number of all antibiotic prescriptions among U.S. children, 2016-2019**

| | All antibiotic prescriptions among U.S. children for any diagnosis | |
|---|---|---|
| **Age stratum** | *Incidence per 1000 person years (95% CI)* | *Annual number, in millions (95% CI)* |
| <2 years | 1,032 (862, 1,226) | 7.8 (6.6, 9.3) |
| 2–4 years | 839 (723, 967) | 10.0 (8.6, 11.6) |
| All children <5 years[a] | 914 (756, 1,095) | 17.9 (14,8, 21.4) |
| 5–17 years | 507 (278, 864) | 27.1 (14.9, 46.2) |
| All children ≤17 years[a] | 627 (515, 757) | 45.8 (37.6, 55.4) |

Abbreviations: CI – confidence interval

[a] Annual numbers of antibiotic prescriptions across age groups may not sum to all children totals due to rounding.

**eTable 14. Sensitivity analysis of vaccine serotype distribution ≥4 years and ≥6 years after PCV13 implementation – serotype distribution among samples with pneumococcal detection[a]**

| | | | Percent of all pneumococcal isolates (95% CI)[a] | | | |
|---|---|---|---|---|---|---|
| **Time period** | **Condition** | **Sample type** | *PCV13[b]* | *PCV15-additional[b]* | *PCV20-additional[b]* | **Studies included** |
| ≥4 years post-PCV13 implementation | AOM | NP | 7.1 (4.1, 11.2) | 6.1 (3.3, 9.7) | 27.4 (21.5, 33.7) | Kaur 2022[14] |
| | ARI | NP | 11.2 (9.3, 13.4) | 4.0 (2.9, 5.4) | 26.2 (23.4, 29.1) | Ben-Shimol 2021;[16] Kaur 2022[14] |
| | AOM | MEF | 18.6 (14.5, 23.4) | 7.3 (4.7, 10.4) | 29.3 (24.3, 34.6) | Ben-Shimol 2021;[26] Kaur 2022;[14] Levy 2019;[22] Marchisio 2017[26] |
| ≥6 years post-PCV13 implementation | AOM | NP | - | - | - | None |
| | ARI | NP | 8.6 (5.4, 12.9) | 4.9 (2.5, 8.2) | 33.6 (27.3. 40.1) | Ben-Shimol 202[16] |
| | AOM | MEF | 11.6 (4.0, 24.0) | 6.7 (1.6, 17.0) | 41.0 (26.1, 56.6) | Ben-Shimol 202[16] |

Abbreviations: PCV – pneumococcal conjugate vaccine; CI – confidence interval; AOM – acute otitis media; NP – nasopharyngeal; ARI – acute respiratory infection; MEF – middle ear fluid

[a] Estimated using Markov-chain Monte Carlo approach based on published estimates from included studies.

[b] Includes vaccine serotypes and corresponding aggregated serotypes/serogroups as detailed in **eTable 6**. PCV-15 additional serotypes (not included in PCV13): 22F, 33F. PCV20-additional serotypes: 8, 10A, 11A, 12F, 15B, 22F, 33F.

### eTable 15. Sensitivity analysis of serotype distribution ≥4 years after PCV13 implementation – proportion of outpatient ARIs attributable to PCV15- and PCV20-additional serotypes[a]

| | **Percent of cases attributable to pneumococcal serotypes (95% CI)** | |
|---|---|---|
| **Condition** | *PCV15-additional* [a] | *PCV20-additional* [a] |
| AOM | 4.1 (1.9, 8.5) | 18.5 (10.5, 33.6) |
| Pneumonia | 2.3 (1.3, 3.9) | 14.9 (9.7, 23.3) |
| Sinusitis | 3.8 (2.3, 6.6) | 25.0 (16.4, 39.0) |

Abbreviations: PCV – pneumococcal conjugate vaccine; ARI – acute respiratory infection; CI – confidence interval; AOM – acute otitis media

[a] Includes vaccine serotypes and corresponding aggregated serotypes/serogroups as detailed in **eTable 6**. PCV-15 additional serotypes (not included in PCV13): 22F, 33F. PCV20-additional serotypes: 8, 10A, 11A, 12F, 15B, 22F, 33F.

### eTable 16. Serotype categorization sensitivity analysis – serotype distribution among samples with pneumococcal detection[a] excluding aggregated serotype and serogroup counts[b]

| | | Percent of all isolates (95% CI)[a] | | |
|---|---|---|---|---|
| **Condition** | **Sample type** | *PCV13*[c] | *PCV15-additional*[c] | *PCV20-additional*[c] |
| ARI | NP | 23.3 (21.5, 25.1) | 3.2 (2.5, 4.1) | 20.0 (18.3, 21.7) |
| AOM | NP | 24.4 (21.1, 27.9) | 4.9 (2.8, 7.7) | 18.3 (14.9, 22.0) |
| AOM | MEF | 35.6 (33.1, 38.2) | 4.6 (3.5, 5.8) | 22.3 (20.1, 24.5) |

Abbreviations: PCV – pneumococcal conjugate vaccine; CI – confidence level; ARI – acute respiratory infection; NP – nasopharyngeal; AOM – acute otitis media; MEF – middle ear fluid

[a] Estimated using Markov chain monte carlo approach based on published estimates from studies in **eTable 1**.

[b] Aggregated serotypes and serogroups considered to be non-vaccine type for Markov chain monte carlo analysis.

[c] Includes vaccine serotypes and corresponding aggregated serotypes/serogroups as detailed in **eTable 6**. PCV-15 additional serotypes (not included in PCV13): 22F, 33F. PCV20-additional serotypes: 8, 10A, 11A, 12F, 15B, 22F, 33F.

**eTable 17. Serotype categorization sensitivity analysis – proportion of outpatient ARIs attributable to PCV15- and PCV20-additional serotypes[a] excluding aggregated serotype and serogroup counts**

| | Percent of cases attributable to pneumococcal serotypes (95% CI) | |
|---|---|---|
| **Condition** | *PCV15-additional* [a] | *PCV15-additional* [a] |
| AOM | 1.0 (0.5, 1.5) | 3.6 (2.7, 4.6) |
| Pneumonia | 0.9 (0.6, 1.4) | 5.4 (3.7, 8.2) |
| Sinusitis | 1.5 (0.9, 2.3) | 9.1 (6.3, 13.7) |

Abbreviations: PCV – pneumococcal conjugate vaccine; CI – confidence interval; AOM – acute otitis media
[a] Includes vaccine serotypes and corresponding aggregated serotypes/serogroups as detailed in **eTable 6**. PCV-15 additional serotypes (not included in PCV13): 22F, 33F. PCV20-additional serotypes: 8, 10A, 11A, 12F, 15B, 22F, 33F.

### eTable 18. Proportion of outpatient ARIs attributable to any serotype in PCV15 and PCV20, including serotypes contained in PCV13[a]

| | Percent of cases attributable to pneumococcal serotypes (95% CI) | |
|---|---|---|
| **Condition** | *All PCV15 serotypes*[b] | *All PCV20 serotypes*[b] |
| AOM | 18.2 (5.6, 32.0) | 21.8 (9.7, 35.3) |
| Pneumonia | 24.3 (22.4, 27.2) | 28.6 (25.4, 33.5) |
| Sinusitis | 19.5 (17.8, 22.0) | 25.8 (22.2, 31.4) |

Abbreviations: ARI – acute respiratory infection; PCV – pneumococcal conjugate vaccine; CI – confidence interval; AOM – acute otitis media

[a] Includes disease prevented by PCV13, residual PCV13 disease, disease due to PCV15-additional serotypes, and disease due to PCV20-additional serotypes. Proportions are computed using a denominator of all disease expected to occur under a scenario without use of any PCV; thus, proportions presented in this table are not directly comparable to those defined using a denominator of current disease burden under the circumstance of ongoing PCV13 use.

[b] Includes serotypes 1, 3, 4, 5, 6A, 6B, 7A, 9V, 14, 18C, 19A, 19F, 23F, 22F and 33F.

[b] Includes serotypes 1, 3, 4, 5, 6A, 6B, 7A, 9V, 14, 18C, 19A, 19F, 23F, 22F, 33F, 8, 10A, 11A, 12F, and 15B.

### eTable 19. Estimated incidence of outpatient visits and antibiotic prescriptions attributable to all serotypes in PCV15 and PCV20,[a] 2016–2019

| | | Estimated incidence per 1000 person-years among U.S. children (95% CI) | | | |
|---|---|---|---|---|---|
| | | *Outpatient visits* | | *Antibiotic prescriptions* | |
| **Age stratum** | **Condition** | All PCV15 serotypes[a] | All PCV20 serotypes[a] | All PCV15 serotypes[a] | All PCV20 serotypes[a] |
| <2 years | | | | | |
| | AOM | 124.3 (33.5, 266.7) | 149.6 (56.3, 297.1) | 88.8 (23.8, 193.0) | 106.9 (39.7, 214.9) |
| | Pneumonia | 12.7 (7.2, 21.3) | 15.0 (8.4, 25.6) | 9.7 (6.6, 14.1) | 11.4 (7.5, 17.1) |
| | Total[b,c] | 137.4 (44.1, 282.6) | 165.0 (68.6, 316.2) | 98.6 (31.8, 204.8) | 118.4 (49.0, 229.2) |
| 2–4 years | | | | | |
| | AOM | 73.1 (19.9, 155.8) | 88.0 (33.5, 173.4) | 66.9 (18.2, 140.2) | 80.4 (30.7, 156.0) |
| | Pneumonia | 8.2 (4.7, 13.4) | 9.6 (5.5, 16.1) | 7.8 (5.5, 11.0) | 9.2 (6.3, 13.3) |
| | Total[b,c] | 81.4 (26.7, 166.1) | 97.8 (41.2, 185.7) | 74.7 (24.6, 150.0) | 89.6 (38.0, 167.7) |
| All children <5 years | | | | | |
| | AOM | 94.4 (25.9, 193.6) | 113.3 (44.0, 214.5) | 76.2 (20.9, 155.9) | 91.6 (3.5, 172.8) |
| | Pneumonia | 10.0 (6.6, 15.0) | 11.8 (7.6, 18.0) | 8.6 (6.1, 11.9) | 10.1 (7.0, 14.5) |
| | Total[b,c] | 104.5 (34.2, 206.0) | 125.3 (53.4, 230.0) | 84.8 (28.1, 166.3) | 101.7 (43.6, 185.7) |
| 5–17 years | | | | | |
| | AOM | 12.9 (3.4, 28.4) | 15.5 (5.7, 31.7) | 13.2 (3.7, 26.9) | 15.9 (6.2, 29.7) |
| | Pneumonia | 2.8 (1.6, 4.7) | 3.3 (1.9, 5.6) | 2.4 (1.7, 3.3) | 2.8 (2.0, 4.0) |
| | Sinusitis | 11.3 (7.4, 16.9) | 15.0 (9.6, 23.4) | 10.4 (6.9, 15.5) | 13.9 (8.9, 21.5) |
| | Total[b] | 27.3 (14.7, 46.4) | 34.2 (19.6, 56.5) | 26.2 (13.9, 43.4) | 32.7 (18.7, 52.7) |
| All children ≤17 years | | | | | |
| | AOM | 34.7 (9.5, 71.3) | 41.7 (16.2, 79.1) | 30.1 (8.3, 60.9) | 36.2 (14.1, 67.4) |
| | Pneumonia | 4.8 (3.2, 6.9) | 5.6 (3.7, 8.4) | 4.7 (3.4, 6.4) | 5.5 (3.9, 7.8) |
| | Sinusitis | 11.2 (7.1, 17.2) | 14.8 (9.1, 23.9) | 9.9 (6.0, 15.9) | 13.2 (7.7, 22.0) |
| | Total[b,d] | 50.9 (22.6, 91.6) | 62.4 (32.0, 106.8) | 45.0 (20.2, 79.6) | 55.1 (28.5, 93.0) |

Abbreviations: CI – confidence interval; PCV – pneumococcal conjugate vaccine; AOM – acute otitis media.

[A] All serotypes include those contained in PCV13 plus PCV15/20-additional serotypes. All serotypes in PCV15: 1, 3, 4, 5, 6A, 6B, 7A, 9V, 14, 18C, 19A, 19F, 23F, 22F, 33F. All serotypes in PCV20: 1, 3, 4, 5, 6A, 6B, 7A, 9V, 14, 18C, 19A, 19F, 23F, 22F, 33F, 8, 10A, 11A, 12F, 15B. See **eMethods 2.3**.

[B] Incidence by condition may not sum to total due to rounding.

[C] Estimates for sinusitis not included in age-stratum totals for <2 years, 2–4 years, and all <5 years. In these strata, sampled visit counts for sinusitis do not meet NAMCS/NHAMCS minimum sample size requirements for valid projection to national estimates.[1] Sinusitis is uncommon in children <5 years due to sinus development.[44,45]

36. Grant LR, Deshpande L, Kimbrough JH, et al. Distribution of serotypes and antimicrobial susceptibility of *Streptococcus pneumoniae* responsible for presumed otitis media among children ≤5 years of age in the United States. Poster presented at: *The 12th International Symposium on Pneumococci and Pneumococcal Diseases (ISPPD-12)*; June 19-23; Toronto, Canada.

37. Candeias C, Alemida, ST, Simões AS, et al. Introduction of PCV13 into the Portuguese National Immunization Plan: Impact on Streptococcus pneumoniae carried by children. Poster presented at: *The 12th International Symposium on Pneumococci and Pneumococcal Diseases (ISPPD-12)*; June 19-23; Toronto, Canada.
